# PURE-MRI: An International Study Assessing Physician Accuracy in Delineating the Prostate and Urethra on Prostate MRI

**DOI:** 10.1101/2025.04.23.25326296

**Authors:** Lily Nguyen, Yuze Song, Anna Dornisch, Madison Baxter, Tristan Barrett, Anders M Dale, Mukesh Harisinghani, Sophia C Kamran, Michael A Liss, Robert T Dess, Daniel JA Margolis, Eric P Weinberg, Tyler M Seibert

**Affiliations:** Department of Radiation Medicine and Applied Sciences, University of California San Diego, La Jolla, CA; University of California San Diego School of Medicine, La Jolla, CA; Department of Electrical and Computer Engineering, University of California San Diego, La Jolla, CA; Department of Radiology, University of Cambridge, Cambridge, United Kingdom; Department of Radiology, University of California San Diego, La Jolla, CA; Department of Radiology, Massachusetts General Hospital, Boston, MA; Department of Radiation Oncology, Massachusetts General Hospital, Harvard Medical School, Boston, MA; Department of Urology, University of California San Diego, La Jolla, CA; Department of Radiation Oncology, University of Michigan, Ann Arbor, MI; Department of Radiology, Cornell University, Ithaca, NY; Department of Clinical Imaging Sciences, University of Rochester Medical Center, Rochester, NY; Department of Bioengineering, University of California San Diego, La Jolla, CA

## Abstract

**Purpose:** Precise delineation of genitourinary structures during prostate cancer (PCa) care is critical to optimize treatment delivery while minimizing toxicity and injury. The Prostate and UREthra on MRI (PURE-MRI) study was an international, prospective study to assess physicians’ accuracy segmenting prostate and urethra on MRI.

**Methods:** Physicians who diagnose or treat PCa were invited to contour prostate and urethra on patient cases using standard *T_2_*-weighted MRI (all planes). We compared these contours to reference consensus segmentations produced by a multidisciplinary panel of experts. We also evaluated performance of a validated prostate auto-segmentation AI tool. Accuracy was assessed with spatial and volumetric analyses.

Mixed effects model was used to evaluate potential factors influencing contour performance.

**Results:** 62 specialists from 11 countries created 114 prostate and 110 urethra contours. Prostate median (min, max) Dice score was 0.92 (0.62, 0.95) for physicians. There was no clear effect of clinical experience or focus. Maximum deviation inside (under-segmentation), maximum deviation beyond expert contour, and mean deviation (per case) from the reference prostate were 3.4 mm (1.0, 12.4), 5.3 mm (2.4, 17.3), and 1.6 mm (0.9, 3.9), respectively. In comparison, prostate auto-segmentation tool results were 0.95 (0.94, 0.96), 3.0 mm, 3.9 mm (3.1, 4.9), and 1.2 mm (1.1, 1.6), respectively. Physician performance was considerably worse for urethra, with Dice score of 0.33 (0.03, 0.69). No urethra AI tool was tested.

**Conclusion:** Physicians contour the prostate on MRI with overall Dice score >0.9, though contours typically had errors >5 mm and sometimes >10 mm. These patterns were observed regardless of clinical experience, specialty, or clinical focus. AI tool performs well enough for clinical use, given comparable accuracy to practicing physicians. In contrast, urethra segmentation on MRI is challenging. More training, better imaging, and/or AI tools may be necessary to achieve consistent, accurate results for the urethra.

## Introduction

Magnetic Resonance Imaging (MRI) is central to diagnosis and local staging of prostate cancer [1–6]. It is also increasingly used for treatment planning. With this increased use, non-radiologists may find more frequent occasion to view and evaluate the prostate and other structures on MRI. One fundamental task is recognizing the gland itself and its boundaries. Radiation oncologists desiring the most precise treatment target for their patients should use MRI, as it generally affords the most accurate view of prostate anatomy. Radiologists routinely measure gland volume and appreciate prostate boundaries to avoid missing intra-prostatic lesions for accurate MRI-ultrasound fusion for targeted biopsy [7]. Urologists often identify the prostate boundary when planning surgery. Physicians from multiple specialties may be interested in the relationship of the prostate to the nearby neurovascular bundle and in assessing proximity of cancer to the urethra [8, 9]. Further, outlining the urethra may be useful for clinicians, as for urologists the length of the membranous urethra is thought to relate to functional outcomes after surgery, and for radiation oncologists they may want to avoid overdosing the urethra [10–12].

Despite the numerous reasons that familiarity with prostate and urethra anatomy on MRI could be clinically relevant, there is little available information as to how well physicians treating prostate cancer can identify these structures. The need for accuracy is arguably greatest for radiation oncologists, who are tasked with delineating the prostate for precise treatment. The FLAME trial provides high-level evidence of improved oncologic outcomes with simultaneous focal radiotherapy boost to MRI-visible disease [13]. Secondary analysis of the same trial suggested urethra dose is associated with toxicity [10]. The urethra is not typically visible on CT scans used for simulation, meaning radiation oncologists need to use MRI. The MIRAGE trial found that reducing the radiotherapy planning treatment volume margin around the prostate from 4 mm to 2 mm significantly reduced toxicity to nearby genitourinary (GU) structures [14], suggesting that errors >2 mm in contouring the prostate itself could be clinically important. Meanwhile, physician variability on contouring the prostate on CT is subject to high variability and relatively large errors [15–17]. However, a recent survey of radiation oncologists highlighted lack of comfort with MRI as a substantial barrier to implementation of focal radiotherapy boost [13], and the ReIGNITE study revealed a lack of familiarity among some radiation oncologists with prostate anatomy on MRI [18, 19].

We sought to evaluate physician accuracy in contouring the prostate and urethra on MRI. Physicians who diagnose and/or treat prostate cancer were invited to participate in the PURE-MRI study. We evaluated inter-physician variability in prostate and urethra contouring, as well as accuracy compared to a multidisciplinary panel’s consensus.

## Methods

To evaluate physician contouring accuracy, we needed prostate MRI scans with reference standards for the prostate and urethra contours developed by experts.

Participants should have had formal clinical training in GU anatomy and experience treating patients with prostate cancer. And we needed to define appropriate and clinically relevant metrics to assess accuracy.

### Patient Cases

Eligible patient cases had undergone prostate multiparametric MRI for known or suspected prostate cancer. A total of 4 patient cases included in this study underwent prostate multiparametric MRI for prostate cancer workup at University of California, San Diego (UCSD) Health between June 2023 to June 2024 (Supplementary Table 1). One case had been chosen because it had been used as a representative case in a previous study of artificial intelligence auto-segmentation tools [20]. Three additional cases were selected among the institutional database for their reasonable image quality and visualized prostate glands. These patients were of ages 67-84 years old and were found to have localized intermediate-risk or high-risk prostate cancer on biopsy.

### Multidisciplinary consensus

Expert-defined contours were obtained to serve as consensus contours to compare with participant and auto-segmented contours. A multidisciplinary, multi-institution team of two genitourinary radiation oncologists (10 years’ experience each) and two genitourinary radiologists (with 12 and 21 years’ experience, respectively) created contours of the prostate in consensus for each of the patients on *T_2_*-weighted MRI [20]. One of the panelists created initial contours on high-resolution axial *T_2_*- weighted images (always viewed in all three planes), which were reviewed in panel meetings, slice by slice. High-resolution coronal and sagittal *T_2_*-weighted acquisitions were available and viewed as needed to complement the axial volume. If any panel member judged the contour deviated > 1 mm from the true prostate boundary, the panel deliberated until unanimous agreement was met. Each slice case was reviewed at least twice. The same review procedures were followed for the urethra. As described previously [20], the panel adopted consensus ground rules to navigate around seminal vesicle tissue and the prostate apex. Likewise, the panel agreed on ground rules for how to contour the urethra. The prostatic urethra and membranous urethra were included. Wherever the urethra was not clearly visible on a given slice (often true at the mid-gland and part of the base of the prostate), the panel used all available anatomic information from the three planes to identify the most likely location. The urethra was always delineated as a continuous structure from the bladder neck to the prostatic apex and membranous urethra, until the most superior axial slice where the penile bulb was fully visible.

### Participant Eligibility

Eligible participants were radiation oncologists, radiologists, or urologists who diagnose and/or treat prostate cancer. Clinical residents or fellows in the above specialties were also eligible if they had completed at least one prostate-focused clinical rotation. Study recruitment materials, communications, and procedures were approved by the UCSD Institutional Review Board (IRB). Participants were recruited via social media, email, and word-of-mouth in the genitourinary oncology community.

### Participant Contours

Participants were randomly assigned 2 of the 4 patient cases on conventional, axial, *T2*-weighted MRI. Contours of prostate and urethra structures were completed using MIM Cloud and MIM Zero Footprint Software (MIM Software, Cleveland, OH). Instructions were provided as a pdf and as a video. We offered one-on-one guidance via teleconference to participants who had difficulty with the software or instructions. Participants were asked to contour the entire prostate. For the urethra, participants were asked to contour prostatic and membranous urethra. All participants were encouraged to complete contours for at least one patient case, with the option to complete more cases if willing.

### Auto-segmentation Artificial Intelligence

For comparison with human contours, a validated prostate auto-segmentation artificial intelligence (AI) tool [20] was used to produce prostate contours on all 4 of the patient cases. No AI tool was evaluated for urethra contours.

### Assessment of accuracy and error

All participant contours were deidentified and compared to the consensus contours. Accuracy of participant prostate contours was assessed using Dice score of the entire prostate (1 indicates perfect overlap; 0 indicates no overlap). AI prostate contours were similarly assessed. To assess deviation from the expert consensus contour boundary, we measured participant under-segmentation via maximum deviation inside (mm) and over-segmentation via maximum deviation outside (mm), respectively. From these metrics, we also calculated the mean deviation error (mm) over the whole prostate boundary. To measure how accurately the superior and inferior extent were defined, we counted slice difference at the prostatic apex and base. This was calculated as the absolute difference in slices between most inferior axial expert contour and participant contour at the apex and similarly with the most superior axial contours at the base of the prostate. Finally, we measured absolute volume difference (%) from expert generated prostate volumes.

To assess accuracy of urethra contours, we compared the clinician generated contours with the expert consensus contours. We analyzed Dice score, volume overlap (%), and Hausdorff distance (mm). First, we reported these metrics along the full length of the urethra (intraprostatic and membranous urethra). We then repeated these analyses for the intraprostatic urethra segmented within the base, middle, and apex, as well as the isolated membranous urethra. We also measured the absolute membranous length difference (mm) compared to expert consensus contours.

Linear mixed effects models were used to evaluate potential factors influencing prostate and urethra contour accuracy. For these models, participants and the MRI patient cases were set as random effects. Fixed effects were set as physician specialty (radiation oncologist, radiologist, or urologist) and clinical experience (still in training, < 5 years, 5-10 years, and >10 years). GU sub-specialization was also set as a fixed effect with the following ordered categories: “Not specialized,” “partially specialized,” or “completely specialized” as self-reported by participants.

## Results

### Cases

A total of 62 physicians from 11 countries participated, representing radiation oncology, radiology, and urology, with a range of sub-specialization and varying years of experience (Table 1). Most of the participants (n=46) completed 2 patient cases, some (n=13) completed only 1 patient case, and few (n=3) completed 3 patient cases. These clinicians produced a total of 224 structure segmentations (114 prostate and 110 urethra contours). 85 (38%) segmentations were done on case 4, 52 (23%) on case 3, 47 (21%) on case 2, and 40 (18%) on case 1.

**Table 1.**
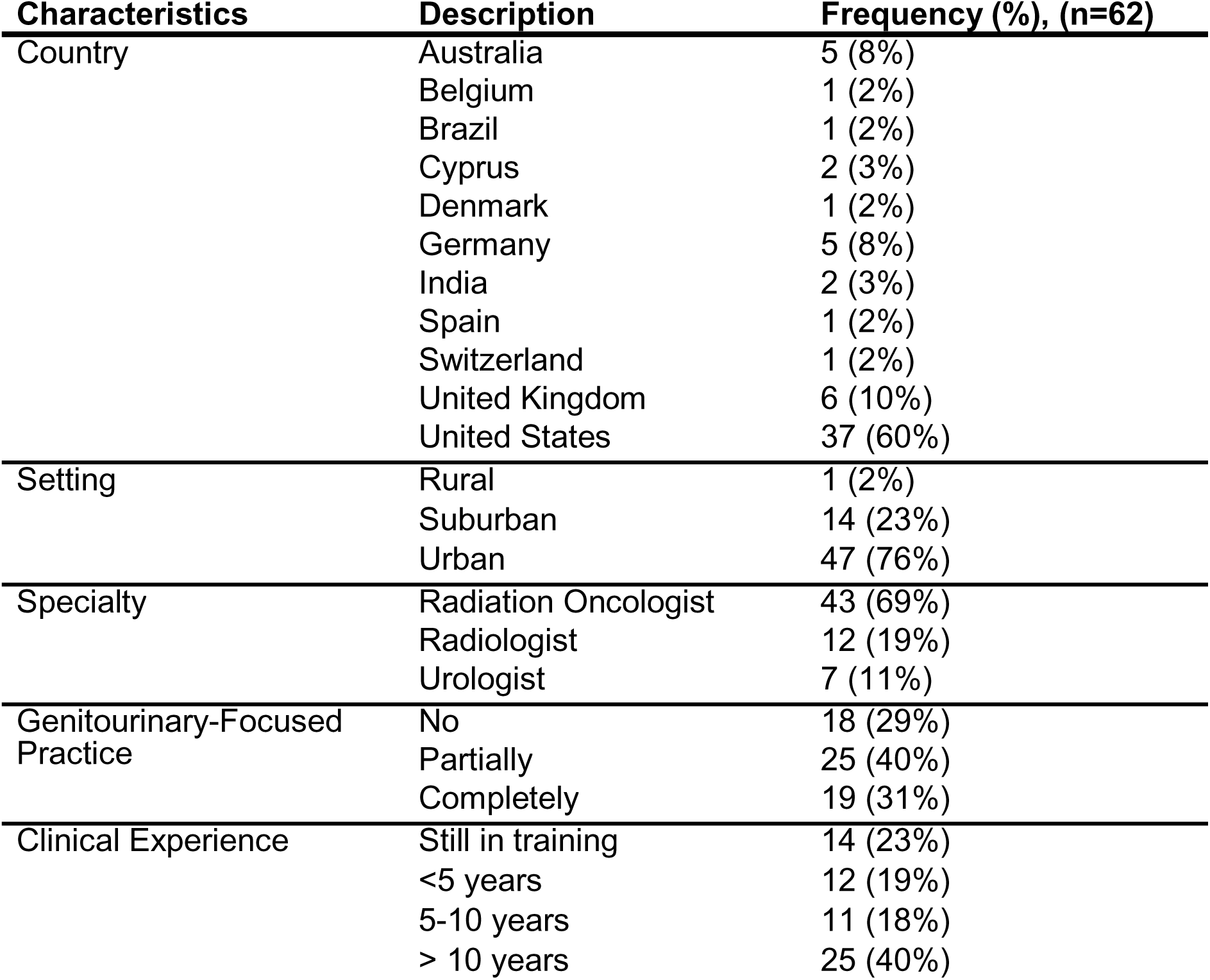
Summary characteristics of participants from survey.

| **Characteristics** | **Description** | **Frequency (%), (n=62)** |
| --- | --- | --- |
| Country | Australia | 5 (8%) |
|  | Belgium | 1 (2%) |
|  | Brazil | 1 (2%) |
|  | Cyprus | 2 (3%) |
|  | Denmark | 1 (2%) |
|  | Germany | 5 (8%) |
|  | India | 2 (3%) |
|  | Spain | 1 (2%) |
|  | Switzerland | 1 (2%) |
|  | United Kingdom | 6 (10%) |
|  | United States | 37 (60%) |
| Setting | Rural | 1 (2%) |
|  | Suburban | 14 (23%) |
|  | Urban | 47 (76%) |
| Specialty | Radiation Oncologist | 43 (69%) |
|  | Radiologist | 12 (19%) |
|  | Urologist | 7 (11%) |
| Genitourinary-Focused Practice | No | 18 (29%) |
|  | Partially | 25 (40%) |
|  | Completely | 19 (31%) |
| Clinical Experience | Still in training | 14 (23%) |
|  | <5 years | 12 (19%) |
|  | 5-10 years | 11 (18%) |
|  | > 10 years | 25 (40%) |

### Assessment of prostate contour accuracy

Overall, participant prostate contours had a median (minimum, maximum) Dice score of 0.92 (0.62, 0.95) (Table 2). Median max deviation error inside and outside for participant contours were 3.4 mm (1.0, 12.4) and 5.3 mm (2.4, 17.3), respectively. At the prostate apex, participant prostate contours had a median slice difference of 1 (0, 5). At the prostate base there was a median slice difference of 0 (0, 5). Participant prostate contour volume differed from expert contour volume by a median of 7.6% (0.1%, 48.2%) (Figure 1C). Among radiation oncologists, Dice score did not appear to vary with years of clinical experience (Figure 2A). Linear mixed effects models did not demonstrate significant, definitive relationships between prostate contour accuracy metrics with specialty, sub-specialization, or clinical experience (Supplementary Tables 2-6).

**Figure 1.**
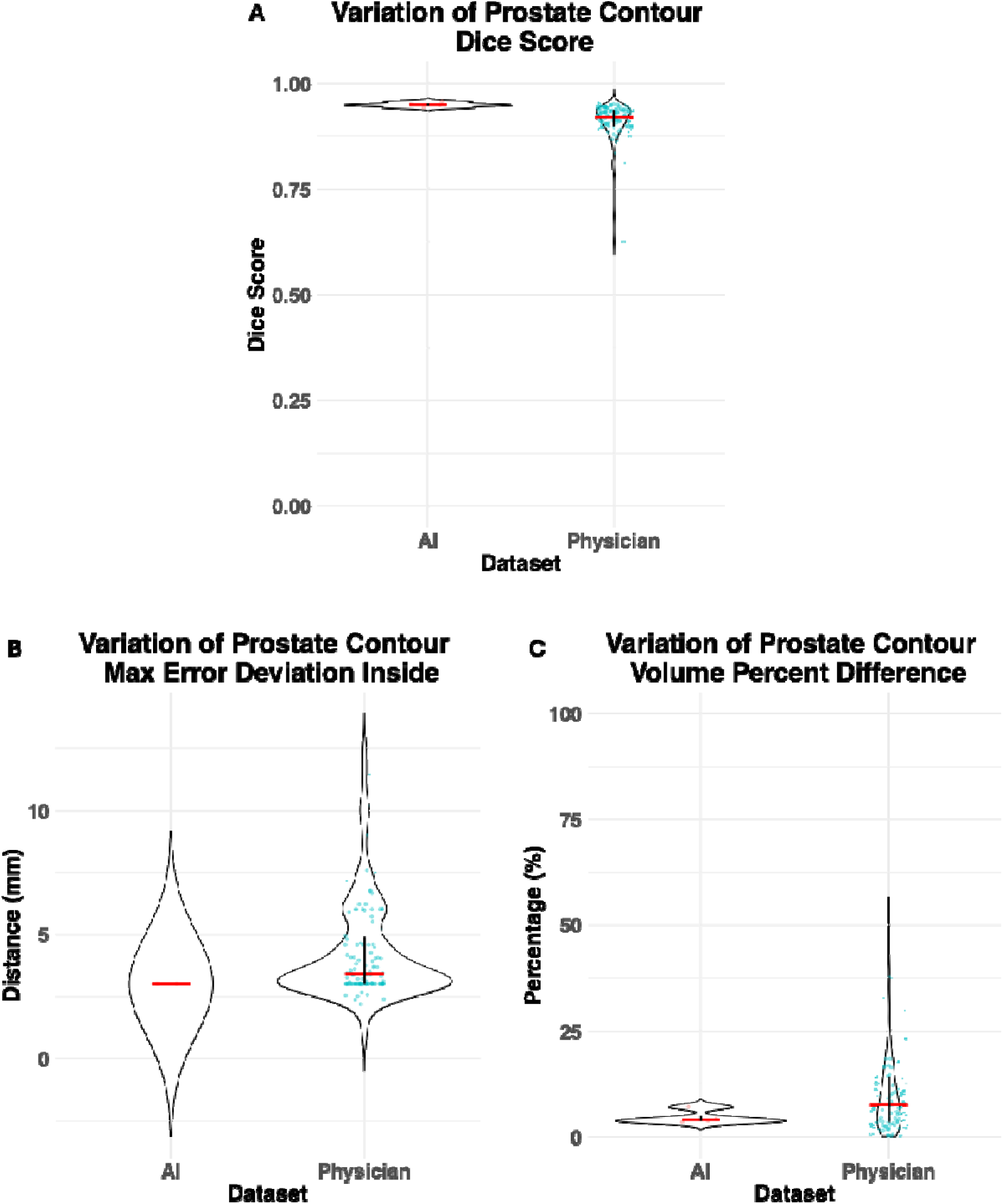
Variation of physician and AI prostate contour accuracy metrics. 1A shows the variation of physician and AI prostate Dice score. 1B shows the variation of max error deviation inside (or under-segmentation from expert consensus contour boundary). 1C shows variation of volume percentage difference from expert consensus contour volume. The median is represented with the red bar and the IQR is represented with the black bar.

**Figure 2.**
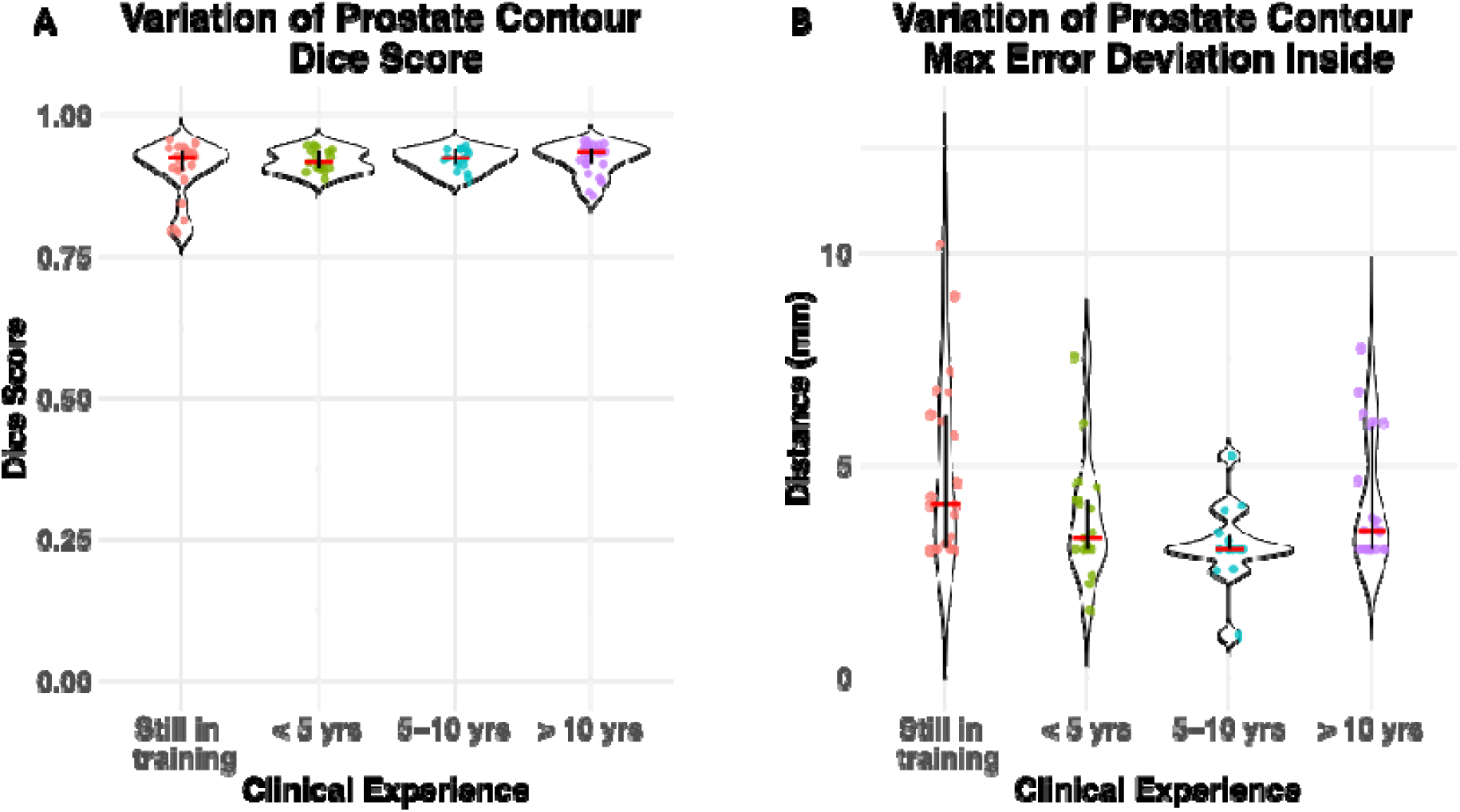
Variation of prostate contour accuracy metrics of radiation oncologists stratified by years of clinical experience. 2A shows the variation of Dice score among radiation oncologists by clinical experience. 2B shows the variation of max error deviation inside (or under-segmentation from expert consensus contour boundary) among radiation oncologists by clinical experience. The median is represented with the red bar and the IQR is represented with the black bar. Yrs= years.

**Table 2.** Physician and AI prostate contour accuracy metrics, reported as median (minimum, maximum).

| Prostate Accuracy Metric |  | Physician Contours,<br>(n=114), median (min,<br>max) | AI Contours, (n=4),<br>median (min, max) |
| --- | --- | --- | --- |
| Dice Score | Whole | 0.92 (0.62, 0.95) | 0.95 (0.94, 0.96) |
| Slice Difference | Apex | 1 (0, 5) | 1.00 (0.75, 1.00) |
|  | Base | 0 (0, 5) | 0 |
| Error Deviation (mm) | Max inside | 3.4 (1.0, 12.4) | 3 |
|  | Max outside | 5.3 (2.4, 17.3) | 3.9 (3.1, 4.9) |
|  | Mean | 1.6 (0.9, 3.9) | 1.2 (1.1, 1.6) |
| Volume Difference (%) | Whole | 7.6 (0.1, 48.2) | 4.0 (3.4, 7.0) |

The AI auto-segmentation tool’s contours had a median Dice score of 0.95 (0.94, 0.96). Other metrics of accuracy for auto-segmentation contours including slice differences at the prostate apex and base, and absolute percent volume differences from consensus contours were within range of participant contour metrics (Figure 1A- C).

### Assessment of urethra contour accuracy

Urethra contours were left unmodified if participants made contiguous segmentations from the base of the bladder to the prostate apex and/or membranous urethra. For participants (n=4) who left some gaps in their urethra contours (e.g., contoured every other slice), the gaps were interpolated using MIM Software’s default tool to allow for consistency in accuracy assessment. Clinician urethra contours had a median Dice score of 0.33 (0.03, 0.69) (Table 3). Urethra contours were also assessed by anatomic region (base, mid, apex, and membranous); contours drawn in the apex region had the highest median Dice score (Supplementary Figure 1A). Median percent volume overlap of clinician urethra contours was 35.7% (3.0%, 95.5%). Similarly, participant urethra contours overlapped the most with expert urethra consensus contours within the apex (Supplementary Figure 1B).

**Table 3.** Physician urethra contour accuracy metrics, reported as median (minimum, maximum). Hausdorff distance was calculated only with urethra structures that included contours drawn in that region. For example, there were 104 urethra structures that had contours that included the base region, and there were only 80 structures that included contours past the apex into the membranous region. Similarly, membranous length difference was calculated only for urethra structures that included contours in the membranous region.

| Urethra Accuracy Metric |  | Physician urethra contours,<br>(n=110), median (min, max) |
| --- | --- | --- |
| Dice Score | Whole | 0.33 (0.03, 0.69) |
|  | Base | 0.23 (0, 0.68) |
|  | Mid | 0.27 (0, 0.68) |
|  | Apex | 0.49 (0.07, 0.79) |
|  | Membranous | 0.30 (0, 0.77) |
| Volume Overlap (%) | Whole | 35.7 (3.0, 95.5) |
|  | Base | 28.7 (0, 100) |
|  | Mid | 35.2 (0, 100) |
|  | Apex | 61.8 (4.1, 100) |
|  | Membranous | 29.5 (0, 100) |
| Hausdorff Distance (mm) | Base | 1.5 (0.6, 2.3), n=104 |
|  | Mid | 1.4 (0.9, 2.2) |
|  | Apex | 1.3 (0.8, 2.2) |
|  | Membranous | 1.3 (0.7, 2.3), n=80 |
| Membranous Length Difference (mm) |  | 5.1 (0, 16.6), n=80 |

Hausdorff distance was calculated in segments relative to prostate region. If a participant left a segment of the prostate empty, we excluded their contour from analysis of that segment. We excluded 6 clinician contours missing urethra segmentation at the base and 30 clinician contours missing membranous urethra segmentation. Hausdorff distance for membranous urethra and intraprostatic urethra at the apex, middle, and base were 1.3 mm (0.7, 2.3), 1.3 mm (0.8, 2.2), 1.4 mm (0.9, 2.2), and 1.5 mm (0.6, 2.3), respectively (Supplementary Figure 1C). Among completed membranous urethra contours (n=80), the median absolute difference in membranous urethra length (compared to the consensus contour) was 5.1 mm (0, 16.6) (Table 3). Linear mixed effects models did not show substantial relationships between participant characteristics and urethral accuracy (Supplementary Tables 7-8).

## Discussion

We analyzed 114 contours of the prostate on MRI by 62 physician participants from 11 countries and found that they generally had excellent average agreement with a multidisciplinary consensus panel (median Dice score 0.92). Encouragingly, these results did not depend heavily on specialty, and inexperienced participants appeared to do about as well as more experienced subspecialists. Despite these overall promising results, though, there were some instances of poor agreement (e.g., Figure 3A) that could result in inadequate treatment of the prostate. Participants also typically had at least one error of over 5 mm and sometimes up to 17 mm, relative to the reference standard. Mean errors across the full prostate boundary were much smaller (median 1-2 mm). Median volume difference was less than 8%.

**Figure 3.**
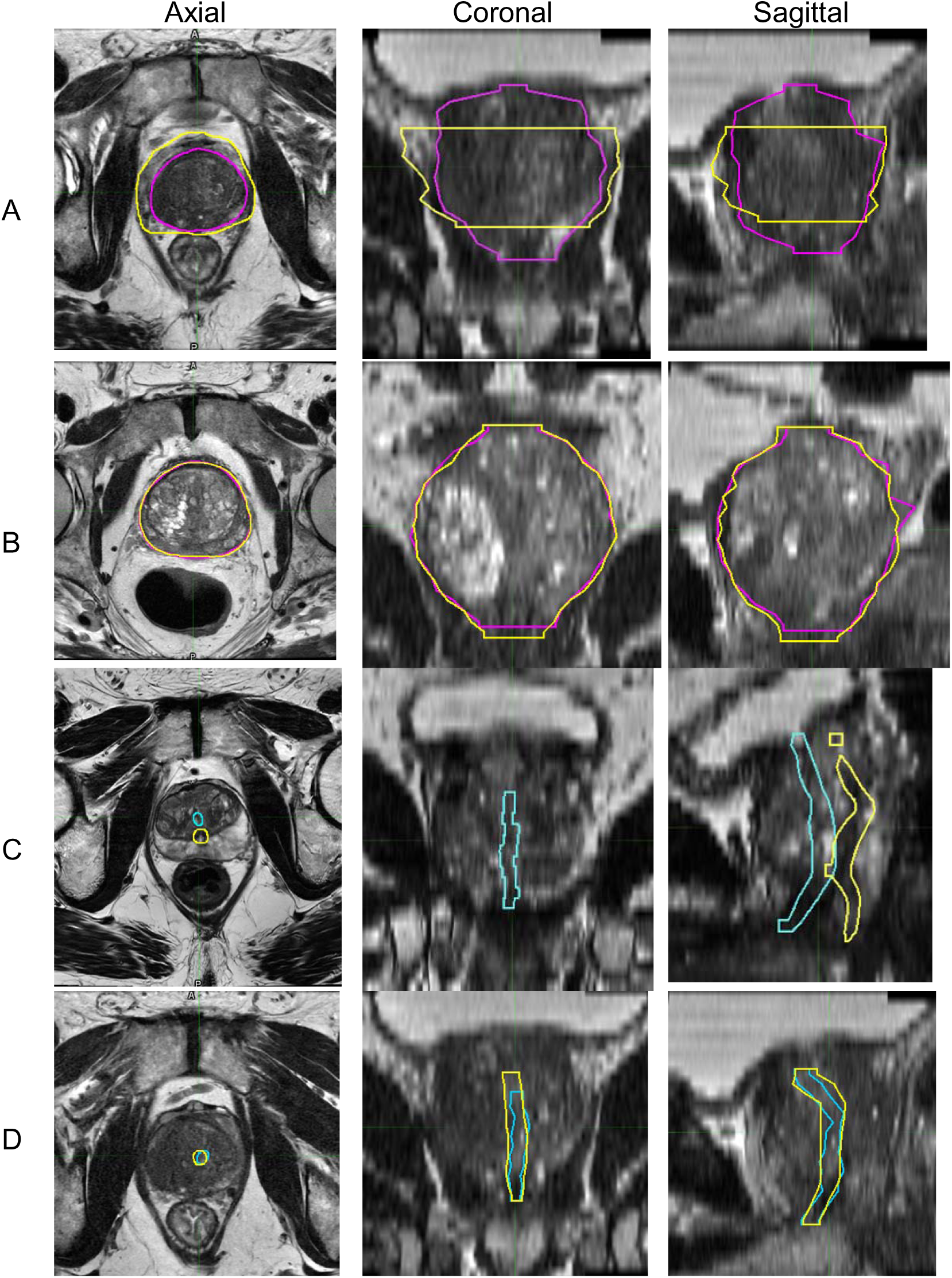
Examples of physician prostate and urethra contours overlayed with expert consensus contours per case on axial, coronal, and sagittal *T2*-weighted MRI. Outlined in yellow were contours that were generated by participant. Outlined in magenta and teal were expert defined prostate and urethra, respectively. 3A is an example of physician prostate contours with poor accuracy as Dice score, mean error deviation, and volume difference were 0.62, 3.9 mm, and 39%, respectively. 3B is an example of high accuracy with Dice score, mean error deviation, and volume difference of 0.95, 1.1 mm, and 4%, respectively. In 3C, physician urethra contours had poor accuracy and overlap with Dice score of 0.03 and volume overlap of 3%. In 3D, these contours had greater Dice score of 0.69 and volume overlap of 85%.

Similar to physician results, an AI auto-segmentation tool performed well for contouring the prostate. The AI tool achieved a Dice score of 0.94-0.96 for each of the four patient cases. The mean error was ≤1.6 mm for each patient case. There were some focal errors where the AI boundary deviated from the panel’s consensus contour, typically by 3-4 mm, but this compares favorably to the human contours. The AI tool had no errors >5 mm, whereas human errors were seen as large as 17 mm. These accurate results are consistent with a previous validation study in a larger number and range of patient cases from multiple institutions and scanners [20]. While the AI tool could be improved, it is already performing at the level of a subspecialist physician.

In line with our findings, previous studies demonstrated excellent agreement between manual contours and gold-standard contours of the prostate on MRI in terms of Dice score [21]. However, though guidelines exist to assist with prostate segmentation accuracy, both intra- and inter-observer variations persist even when using the same imaging dataset [22, 23]. As a result, prostate manual segmentation comes with substantial uncertainty. In parallel, with the rapid emergence of AI applications and widespread implementation of AI algorithms into medical imaging, auto-segmentation tools are rising. Other studies have also shown good performance of AI tools for the prostate, with a growing body of evidence demonstrating promise for integration of auto-segmentation tools to improve efficiency of clinical workflow [20, 24–27]. Such tools would also reduce the intra- and inter-physician variability reported previously [22, 23].

On the other hand, the urethra poses as a larger challenge as physicians struggled to delineate the urethra. This is relevant, as there is substantial international interest in sparing the urethra in hopes of reducing treatment toxicity [28–30]. Dice scores were very low, with a median of 0.33. The worst urethra contour barely touched the reference urethra (Dice score 0.03; Figure 3C), and even the best physician contour only achieved a modest Dice score of 0.69. One possibility is participants could have achieved poor Dice scores because they were conservatively over-contouring to encompass the urethra broadly. This appears not to be the case, though, as the median percentage of the consensus urethra encompassed by participants’ contours was less than 36%, with all segments except the apex seeing at least one physician give 0% coverage. Regions of the prostatic urethra near the middle and base of the prostate had lower Dice scores which could be due to the anatomic path of the urethra being more oblique as it passes through the prostate superiorly, which limits axial visualization. These results raise significant questions regarding the reliability of physician urethra contours. Studies reporting analysis of radiation dose-dependent toxicity are likewise suspect without careful central review of urethra contours.

Given the low bar of current human performance, an AI tool could be very useful for research (normal-tissue complication probability analyses) and even for clinical practice. Currently, we are working to develop a model that can achieve high accuracy and have invited several companies with existing tools to allow us to evaluate them.

These studies are left for future work. While we await validation and availability of suitable AI tools, we suggest more educational initiatives are warranted, perhaps through the major radiation oncology and radiology society meetings. There are no definitive guidelines for delineation of the urethra [31], and it is a challenging task. The multidisciplinary panel for the present study notes that the membranous and apical urethra are typically visible. The bladder neck is also typically visible, especially on sagittal views. The intraprostatic urethra in the mid-gland and base can be substantially more difficult and is sometimes not visible. The urethra generally sits between the two halves of the central gland, though benign prostatic hyperplasia nodules can cause significant deviation from midline in many patients. Finally, visible parts of the urethra must be connected to each other, allowing for some sensible interpolation. We are publishing here the panel consensus contours for all slices from one example patient case as an educational resource (Supplementary Figure 2).

Other possibilities for delineating the urethra include voiding cystourethrography, retrograde urethrography, or transrectal ultrasonography [31]. Placement of a catheter will make the urethra readily visible but is invasive and distorts the urethral anatomy such that accuracy might require placement of the catheter for every radiation treatment, for example [32, 33]. Simpler surrogates for the urethra have shown promise and could be evaluated further [34]. Some centers use a 3D *T_2_*-weighted acquisition (analogous to 3D Fluid-Attenuated Inversion Recovery sometimes used for brain MRI) to help find the urethra. The multidisciplinary panel here evaluated images from several centers and concluded that there was not a clear benefit, as the axial reconstructions tend to be blurrier than 2D axial series due to the increased susceptibility of 3D acquisitions to motion artifact. Still, this may prove to be an effective strategy with more investigation. At least one study suggests micturition just prior to prostate MRI makes the urethra more visible [35]. This could be a viable approach, though most diagnostic imaging centers already suggest patients use the restroom prior to MRI for practical reasons (and MRI simulation for prostate radiotherapy would often entail a full bladder).

Our study has some limitations. First, our study included only four patient cases, as we favored being able to directly compare multiple contours on the same case. Also, in isolation, Dice scores and other methods to evaluate spatial and volumetric structures lack specificity to understand if the errors measured are clinically significant [25].

Furthermore, there is hesitation to establish cut-off threshold values for these metrics as qualitative assessments by physicians may remain a valuable component [25]. Second, while outside the scope of this study, clinical use of prostate and urethra segmentation for radiotherapy or surgery has additional considerations, such as need for registration between imaging modalities and physiological changes of the prostate position between scan and treatment. Further, we acknowledge that some anatomical variations or disease states may make outlining more challenging (e.g. peri-prostatic veins, large volume disease with extracapsular extension, utricle/Wolffian duct cysts, atrophic seminal vesicles, etc).

## Conclusion

Regardless of clinical experience, specialty, or clinical focus, physicians were able to contour the prostate on MRI with typically (but not universally) excellent Dice scores (>0.90). An AI prostate segmentation tool performed at least as well as human physician participants. Contouring the urethra on MRI is more challenging, calling into question the feasibility of clinicians to adopt more urethral-sparing techniques for radiotherapy without more widespread education or development of more robust techniques.

## Data Availability

All data produced in the present study are available upon reasonable request to the authors

## Notes

### Competing Interest Statement

AMD is a founder of and holds equity interest in CorTechs Labs and serves on its scientific advisory board. He is also a member of the Scientific Advisory Board of Healthlytix and receives research funding from General Electric Healthcare (GEHC). MH reports having received research funding from Perspectum. SK reports honoria from DAVA Oncology and Varian. She also receives funding from NRG Oncology and Prostate Cancer Foundation. She also serves leadership roles in American Board of Radiology, ASCO, and MGH. ML is the founder of Oncobiomix. RTD had served on the advisory board from Janssen Pharmaceuticals and he is currently co-director of Michigan Radiation Oncology Quality Consortium. DJAM reports having had a consultant role for Promaxo, Inc. and Guebert, Inc. TMS reports honoraria from Varian Medical Systems, WebMD, MJH Life Sciences, GE Healthcare, Blue Earth Diagnostics, and Janssen; he has an equity interest in CorTechs Labs, Inc. and serves on its Scientific Advisory Board; he receives research funding from GE Healthcare and Blue Earth Diagnostics, as well as in-kind research support from Quibim, Inc., both through the University of California San Diego. These companies might potentially benefit from the research results. The terms of this arrangement have been reviewed and approved by the University of California San Diego in accordance with its conflict-of-interest policies.

### Funding Statement

National Institutes of Health, Grant/Award Numbers: NIH/NIBIB K08EB026503, NIHUL1TR000100;
American Society for Radiation Oncology, the Prostate Cancer Foundation: PCF20YOUN01;
Department of Defense, Grant/Award Number: DOD/CDMRPPC220278;

### Author Declarations

IRB of University of California San Diego gave ethical approval for this work

### Summary of Updates

Changes were made to the order of figures and tables referred in the manuscript

