## Supplementary Material for "PURE-MRI: An International Study Assessing Physician Accuracy in Delineating the Prostate and Urethra on Prostate MRI"

| Scanner Characteristics | Axial *T2* | Sagittal *T2* | Coronal *T2* |
| --- | --- | --- | --- |
| Pulse Sequence | FSE | FSE | FSE |
| TR (ms) | 3276-3615* | 2662-4684* | 2662-3357* |
| TE (ms) | 98-172* | 98-173* | 98-167* |
| FOV (mm) | 160 x 160, 180 x 180 | 160 x 160, 180 x 180 | 160 x 160, 180 x 180 |
| Matrix (resampled dimensions) | 360 x 224, 400 x 256 | 360 x 224, 384 x 384 | 360 x 224, 400 x 400 |
| Slices | 32 | 26 | 26 |
| Slice Thickness (mm) | 3 | 3 | 3 |

** TR and TE reported as minimum and maximum range, as these measurements varied for each patient case.*

Supplementary Table 1. Acquisition parameters for *T2*-weighted MRI sequences for patient cases used in this study. For FOV, there were two sets listed as one of the patient cases had different matrix dimensions for each plane; the other three patient cases had the same matrix dimensions of 360 x 224 for all planes. TR: repetition time. TE: echo time. FOV: field-of-view. FSE: fast spin echo.

| Fixed Effects Coefficients | β (95% CI) | SE | t stat | p-value |
| --- | --- | --- | --- | --- |
| Intercept | 0.89 (0.86, .92) | 0.02 | 56.06 | < 2e-16 |
| Radiologist | -0.01 (-0.04, 0.01) | 0.01 | -0.80 | 0.42 |
| Urologist | -0.05 (-0.08, -0.02) | 0.02 | -2.78 | 0.01 |
| GU-Focused Practice | -0.01 (-0.03, 0.01) | 0.01 | -0.56 | 0.58 |
| Clinical Experience <5 years | 0.03 (0.00, 0.07) | 0.02 | 1.94 | 0.06 |
| Clinical Experience 5-10 years | 0.02 (-0.02, 0.05) | 0.02 | 0.97 | 0.34 |
| Clinical Experience >10 years | 0.02 (0.00, 0.05) | 0.01 | 1.55 | 0.13 |

Supplementary Table 2. Linear mixed effects model for prostate Dice score. Linear mixed effects model evaluation of prostate Dice score and random effects of MRI case assignment and participant. Fixed effect coefficients included physician specialty with radiation oncologist as the reference. For practice specialization, “not GU-focused” was set as the reference with partially and completely GU-focused specialization set in ordered, categories. For years of clinical experience, “still in training” was set as the reference group. GU= genitourinary. β = coefficient of the model. SE = standard error for model coefficient.

| Fixed Effects Coefficients | β (95% CI) | SE | t stat | p-value |
| --- | --- | --- | --- | --- |
| Intercept | 6.2 (4.5, 8.0) | 0.9 | 6.97 | 1.27e-05 |
| Radiologist | 0.3 (-1.1, 1.7) | 0.8 | 0.37 | 0.71 |
| Urologist | 2.0 (0, 3.8) | 1.0 | 1.93 | 0.06 |
| GU-Focused Practice | 1.1 (0, 2.3) | 0.6 | 1.78 | 0.08 |
| Clinical Experience <5 years | -0.8 (-2.7, 1.0) | 1.0 | -0.83 | 0.41 |
| Clinical Experience 5-10 years | 0 (-1.8, 1.9) | 1.0 | 0.01 | 0.99 |
| Clinical Experience >10 years | -0.6 (-2.2, 1.0) | 0.9 | -0.71 | 0.48 |

Supplementary Table 3. Linear mixed effects model for prostate max error deviation outside (mm). Linear mixed effects model evaluation of prostate max error deviation outside (mm) beyond expert consensus contour and random effects of MRI case assignment and participant. Fixed effect coefficients included physician specialty with radiation oncologist as the reference. For practice specialization, “not GU-focused” was set as the reference with partially and completely GU-focused specialization set in ordered, categories. For years of clinical experience, “still in training” was set as the reference group. GU= genitourinary. β = coefficient of the model. SE = standard error for model coefficient.

| Fixed Effects Coefficients | β (95% CI) | SE | t stat | p-value |
| --- | --- | --- | --- | --- |
| Intercept | 4.6 (3.6, 5.7) | 0.6 | 8.31 | 6.29e-11 |
| Radiologist | 1.1 (0, 2.2) | 0.6 | 1.86 | 0.07 |
| Urologist | 0.9 (-0.6, 2.4) | 0.8 | 1.12 | 0.27 |
| GU-Focused Practice | -0.8 (-1.7, 0.1) | 0.5 | -1.58 | 0.12 |
| Clinical Experience <5 years | -1.0 (-2.5, 0.4) | 0.8 | -1.35 | 0.18 |
| Clinical Experience 5-10 years | -0.9 (-2.3, 0.6) | 0.8 | -1.13 | 0.26 |
| Clinical Experience >10 years | -0.7 (-1.9, 0.6) | 0.7 | -0.98 | 0.33 |

Supplementary Table 4. Linear mixed effects model for prostate max error deviation inside (mm). Linear mixed effects model evaluation of prostate max error deviation inside (mm), or under-segmentation from expert consensus contour boundary, and random effects of MRI case assignment and participant. Fixed effect coefficients included physician specialty with radiation oncologist as the reference. For practice specialization, “not GU-focused” was set as the reference with partially and completely GU-focused specialization set in ordered, categories. For years of clinical experience, “still in training” was set as the reference group. GU= genitourinary. β = coefficient of the model. SE = standard error for model coefficient.

| Fixed Effects Coefficients | β (95% CI) | SE | t stat | p-value |
| --- | --- | --- | --- | --- |
| Intercept | 2.0 (1.5, 2.4) | 0.2 | 8.36 | 1.37e-05 |
| Radiologist | 0.2 (-0.1, 0.6) | 0.2 | 1.23 | 0.23 |
| Urologist | 0.4 (0, 0.9) | 0.2 | 1.76 | 0.08 |
| GU-Focused Practice | 0.2 (0, 0.5) | 0.1 | 1.38 | 0.17 |
| Clinical Experience <5 years | -0.4 (-0.9, 0) | 0.2 | -1.83 | 0.07 |
| Clinical Experience 5-10 years | -0.2 (-0.7, 0.2) | 0.2 | -0.97 | 0.34 |
| Clinical Experience >10 years | -0.3 (-0.6, 0.1) | 0.2 | -1.25 | 0.22 |

Supplementary Table 5. Linear mixed effects model for prostate mean error deviation (mm). Linear mixed effects model evaluation of prostate mean error deviation (mm) from expert consensus contour and random effects of MRI case assignment and participant. Fixed effect coefficients included physician specialty with radiation oncologist as the reference. For practice specialization, “not GU-focused” was set as the reference with partially and completely GU-focused specialization set in ordered, categories. For years of clinical experience, “still in training” was set as the reference group. GU= genitourinary. β = coefficient of the model. SE = standard error for model coefficient.

| Fixed Effects Coefficients | β (95% CI) | SE | t stat | p-value |
| --- | --- | --- | --- | --- |
| Intercept | 13.7 (8.8, 18.7) | 2.7 | 5.17 | 2.38e-05 |
| Radiologist | -0.5 (-5.5, 4.5) | 2.6 | -0.20 | 0.84 |
| Urologist | 6.4 (-0.3, 12.9) | 3.5 | 1.81 | 0.08 |
| GU-Focused Practice | 2.6 (-1.3, 6.7) | 2.1 | 1.23 | 0.22 |
| Clinical Experience <5 years | -4.7 (-11.1, 1.9) | 3.5 | -1.35 | 0.18 |
| Clinical Experience 5-10 years | -4.7 (-11.0, 1.8) | 3.4 | -1.38 | 0.17 |
| Clinical Experience >10 years | -4.8 (-10.4, 0.8) | 3.0 | -1.62 | 0.11 |

Supplementary Table 6. Linear mixed effects model for prostate volume percentage difference. Linear mixed effects model evaluation of prostate volume percentage difference from consensus contour and random effects of MRI case assignment and participant. Fixed effect coefficients included physician specialty with radiation oncologist as the reference. For practice specialization, “not GU-focused” was set as the reference with partially and completely GU-focused specialization set in ordered, categories. For years of clinical experience, “still in training” was set as the reference group. GU= genitourinary. β = coefficient of the model. SE = standard error for model coefficient.

| Fixed Effects Coefficients | β (95% CI) | SE | t stat | p-value |
| --- | --- | --- | --- | --- |
| Intercept | 0.37 (0.30, 0.45) | 0.04 | 9.92 | 4.88e-09 |
| Radiologist | -0.11 (-0.18, -0.05) | 0.04 | -3.25 | 0.002 |
| Urologist | -0.14 (-0.23, -0.05) | 0.05 | -2.76 | 0.01 |
| GU-Focused Practice | 0.04 (-0.01, 0.10) | 0.03 | 1.43 | 0.16 |
| Clinical Experience <5 years | 0.02 (-0.07, 0.10) | 0.05 | 0.33 | 0.74 |
| Clinical Experience 5-10 years | 0.01 (-0.07, 0.10) | 0.05 | 0.26 | 0.80 |
| Clinical Experience >10 years | -0.01 (-0.08, 0.07) | 0.04 | -0.22 | 0.83 |

Supplementary Table 7. Linear mixed effects model for urethra Dice score. Linear mixed effects model evaluation of urethra Dice score and random effects of MRI case assignment and participant. Fixed effect coefficients included physician specialty with radiation oncologist as the reference. For practice specialization, “not GU-focused” was set as the reference with partially and completely GU-focused specialization set in ordered, categories. For years of clinical experience, “still in training” was set as the reference group. Results suggest profession has a potential effect on poorer Dice scores (-0.11) for radiologists with a p-value of 0.002. However, this is a not a robust effect, as only 19% (n=12) of the participants were radiologists. GU= genitourinary. β = coefficient of the model. SE = standard error for model coefficient.

| Fixed Effects Coefficients | β (95% CI) | SE | t stat | p-value |
| --- | --- | --- | --- | --- |
| Intercept | 37.1 (13.7, 22.7) | 6.7 | 5.50 | 1.26e-06 |
| Radiologist | -12.0 (-25.9, 2.0) | 7.6 | -1.58 | 0.12 |
| Urologist | 7.5 (-12.0, -27.2) | 10.6 | 0.71 | 0.48 |
| GU-Focused Practice | 5.0 (-6.3, 16.3) | 6.1 | 0.82 | 0.42 |
| Clinical Experience <5 years | 5.5 (-12.5, 23.5) | 9.8 | 0.57 | 0.58 |
| Clinical Experience 5-10 years | 12.7 (-5.5, 30.7) | 9.8 | 1.29 | 0.20 |
| Clinical Experience >10 years | 4.5 (-11.2, 20.1) | 8.5 | 0.53 | 0.60 |

Supplementary Table 8. Linear mixed effects model for urethra volume overlap percentage. Linear mixed effects model evaluation of urethra volume overlap percentage with expert consensus contours and random effects of MRI case assignment and participant. Fixed effect coefficients included physician specialty with radiation oncologist as the reference. For practice specialization, “not GU-focused” was set as the reference with partially and completely GU-focused specialization set in ordered, categories. For years of clinical experience, “still in training” was set as the reference group. GU= genitourinary. β = coefficient of the model. SE = standard error for model coefficient.

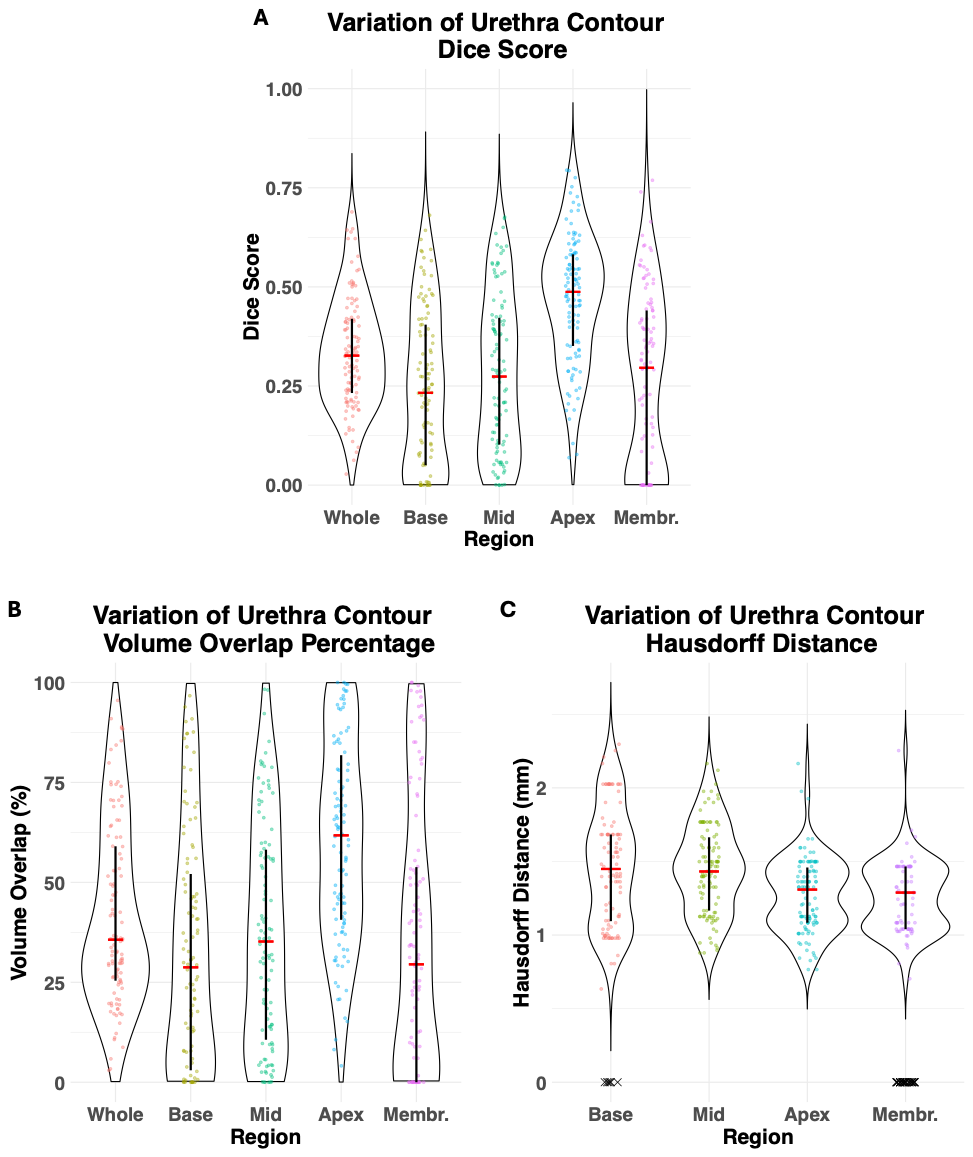

Supplementary Figure 1. Variation of urethra contour accuracy metrics. 1A shows variation of urethra contour Dice score relative to anatomical prostate region. 1B shows variation of physician urethra contour volume overlap percentage with expert consensus contour relative to anatomical prostate region. 1C shows urethra contour Hausdorff distance per anatomical prostate region. The x’s represent excluded urethra contours, as these physician contours did not include segmentations in these specific anatomic regions. The median is represented with the red bar and the interquartile range is represented with the black bar. Membr.= Membranous.

| 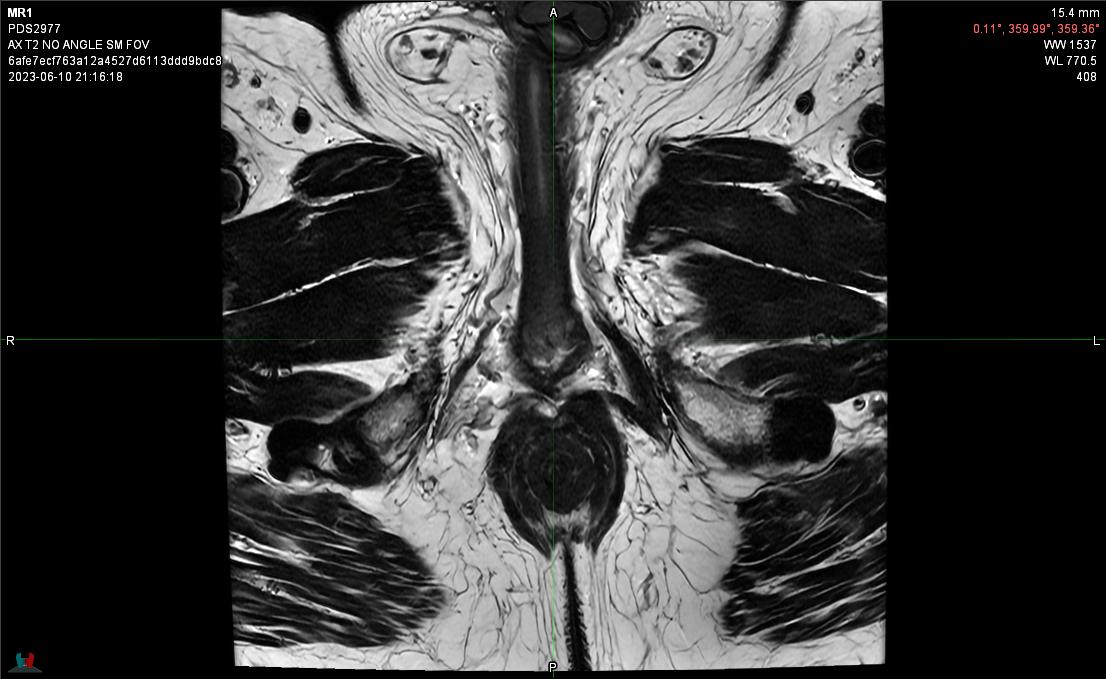  Axial View Slice 1 | 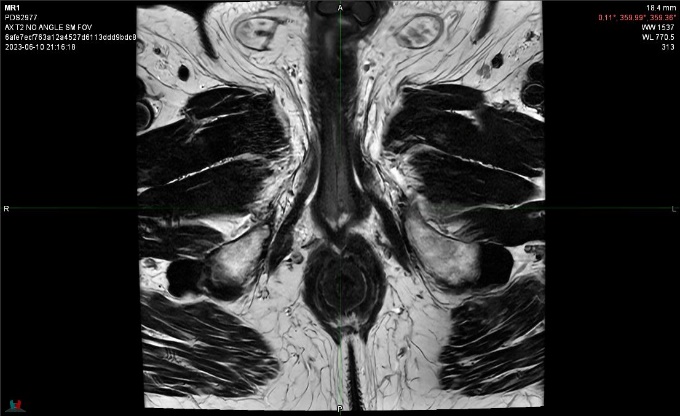  Axial View Slice 2 |
| --- | --- |
| 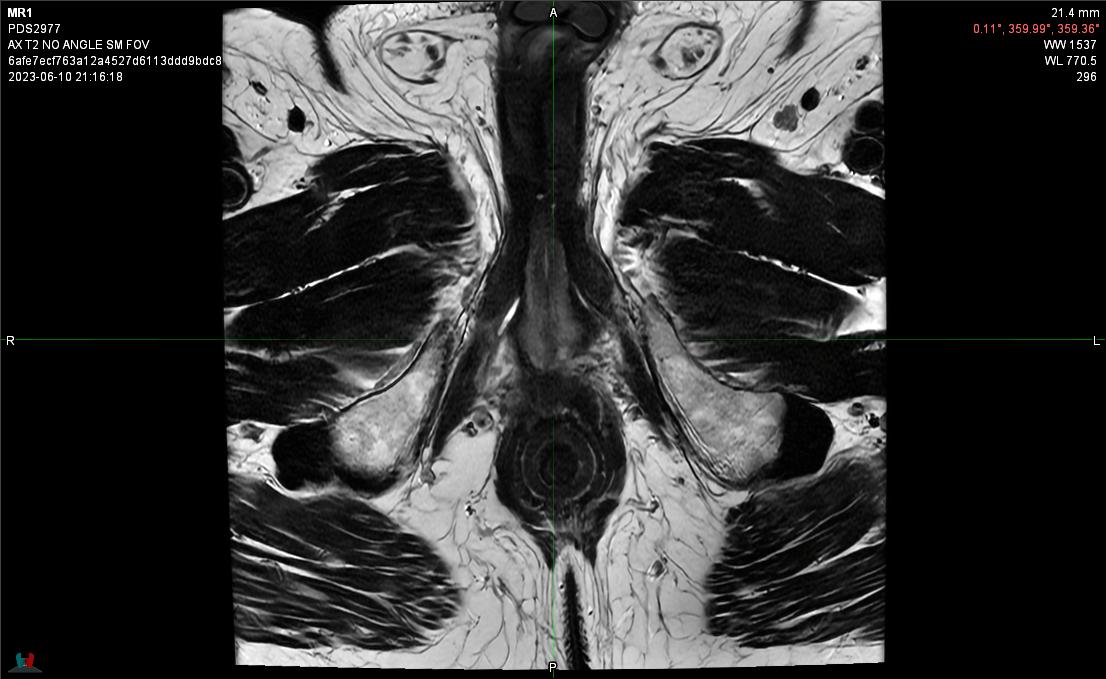  Axial View Slice 3 | 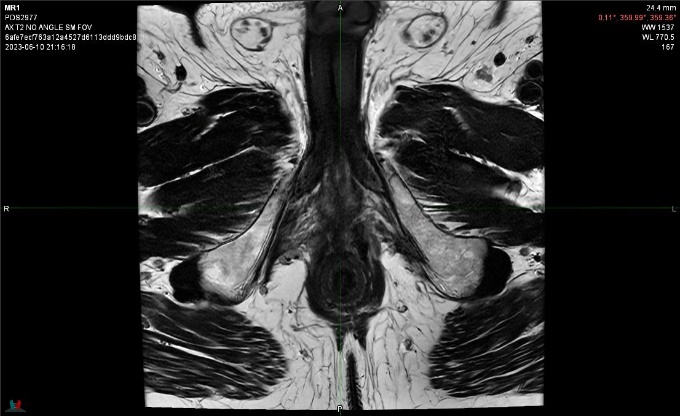  Axial View Slice 4 |
| 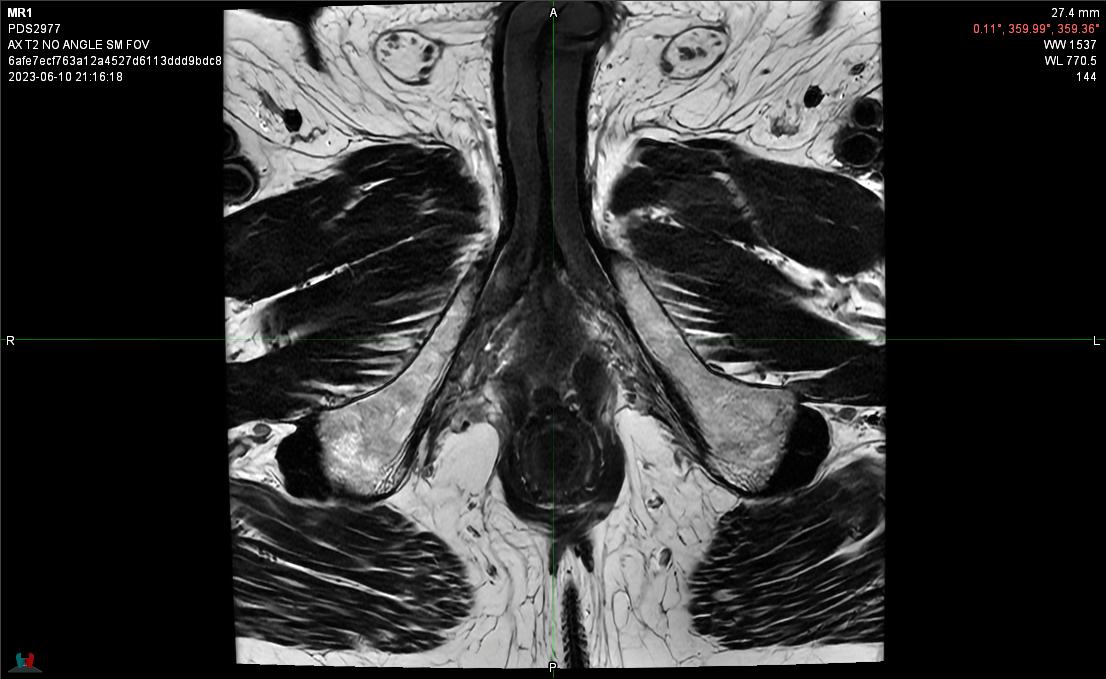  Axial View Slice 5 | 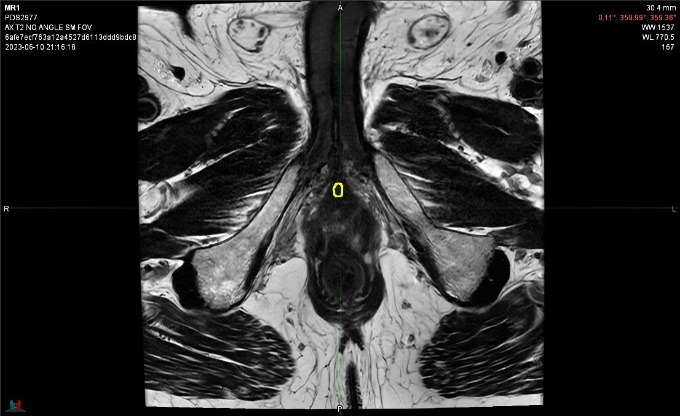  Axial View Slice 6 |
| 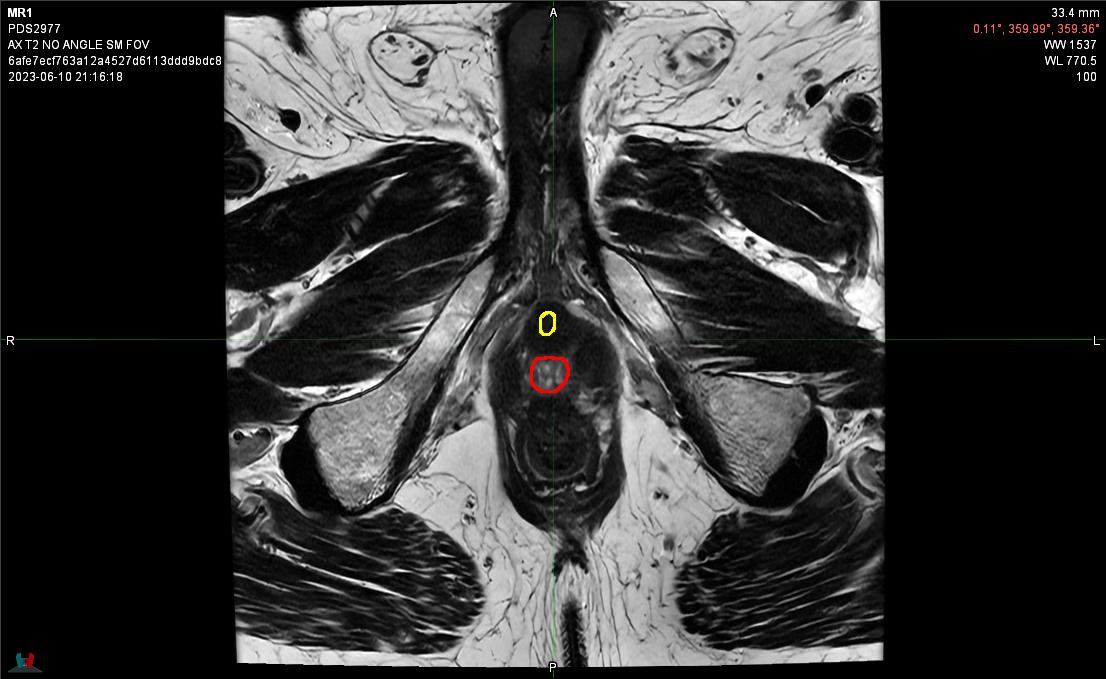  Axial View Slice 7 | 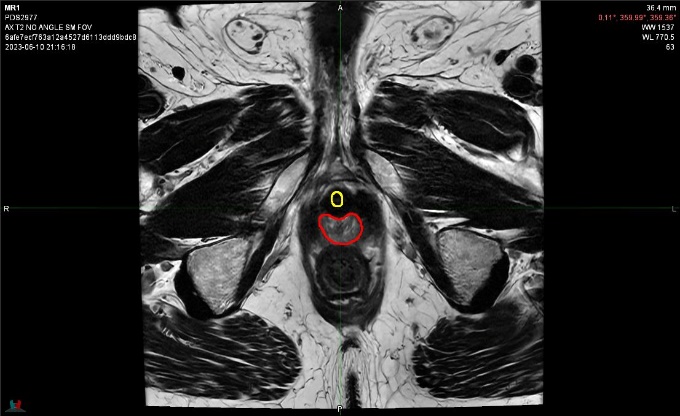  Axial View Slice 8 |
| 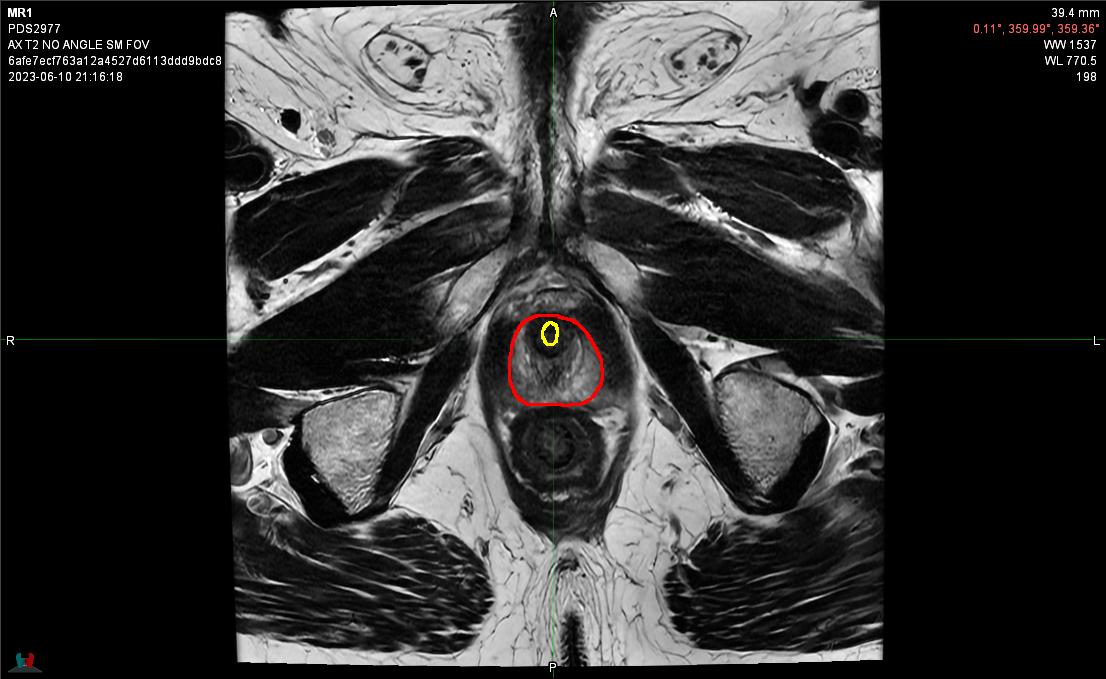  Axial View Slice 9 | 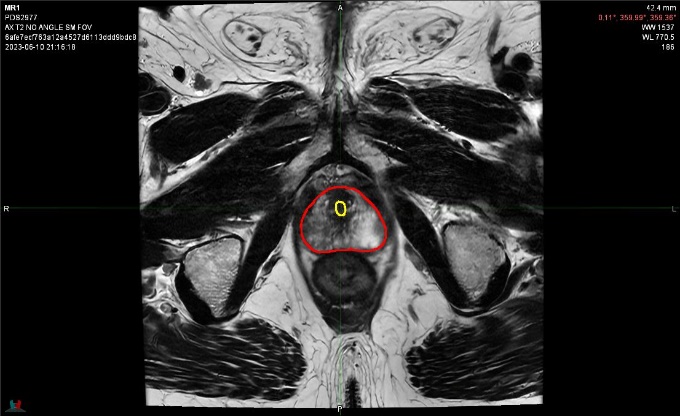  Axial View Slice 10 |
| 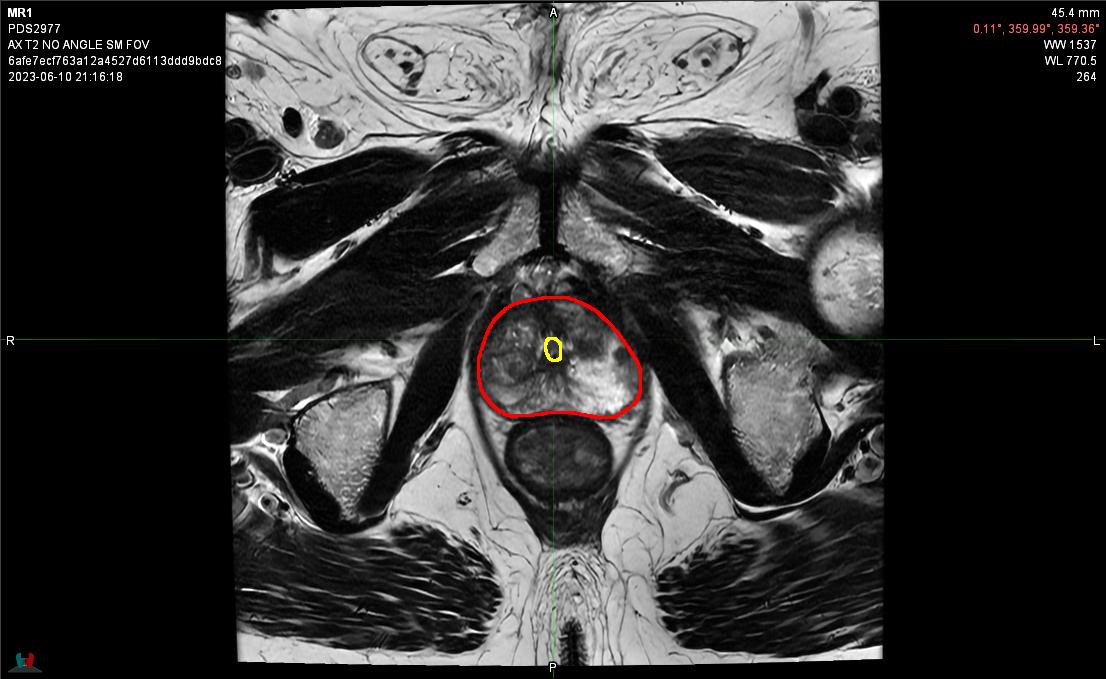  Axial View Slice 11 | 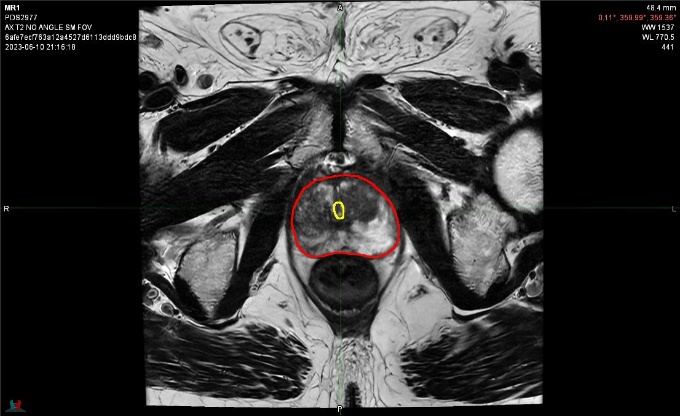  Axial View Slice 12 |
| 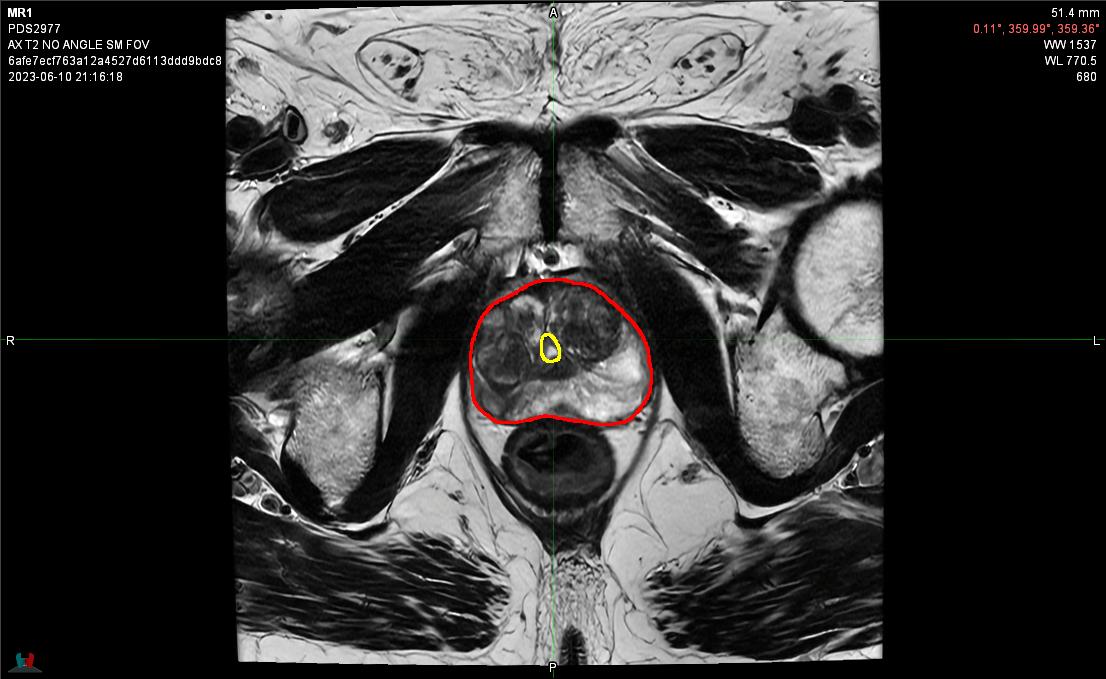  Axial View Slice 13 | 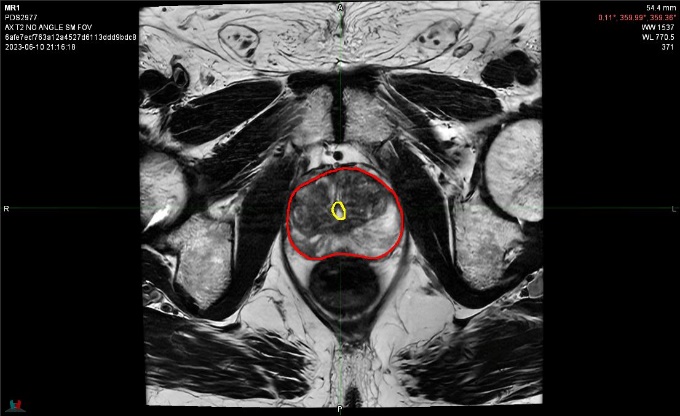  Axial View Slice 14 |
| 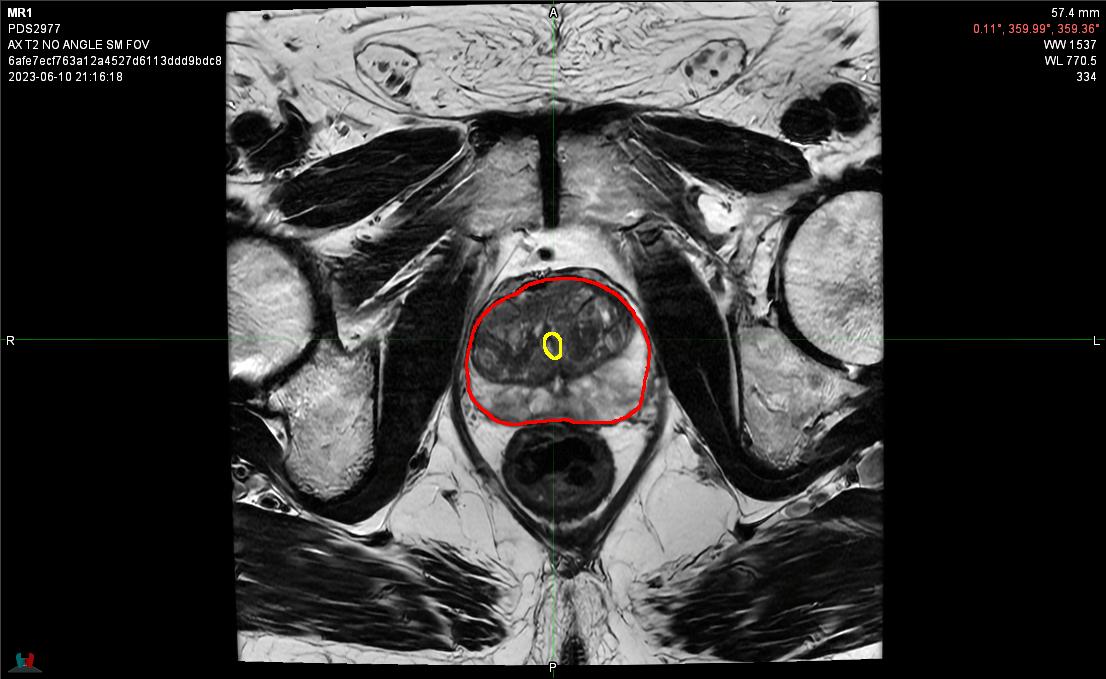  Axial View Slice 15 | 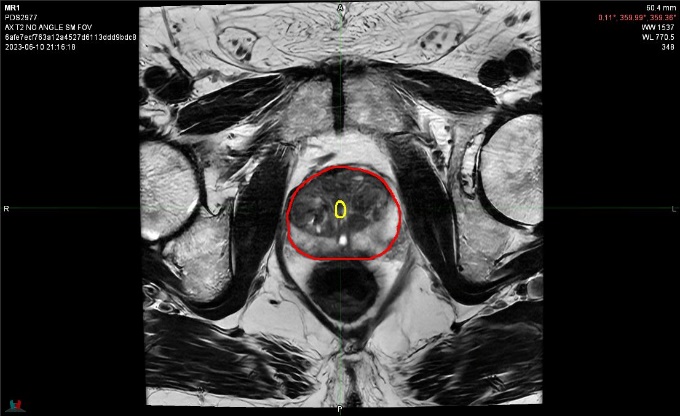  Axial View Slice 16 |
| 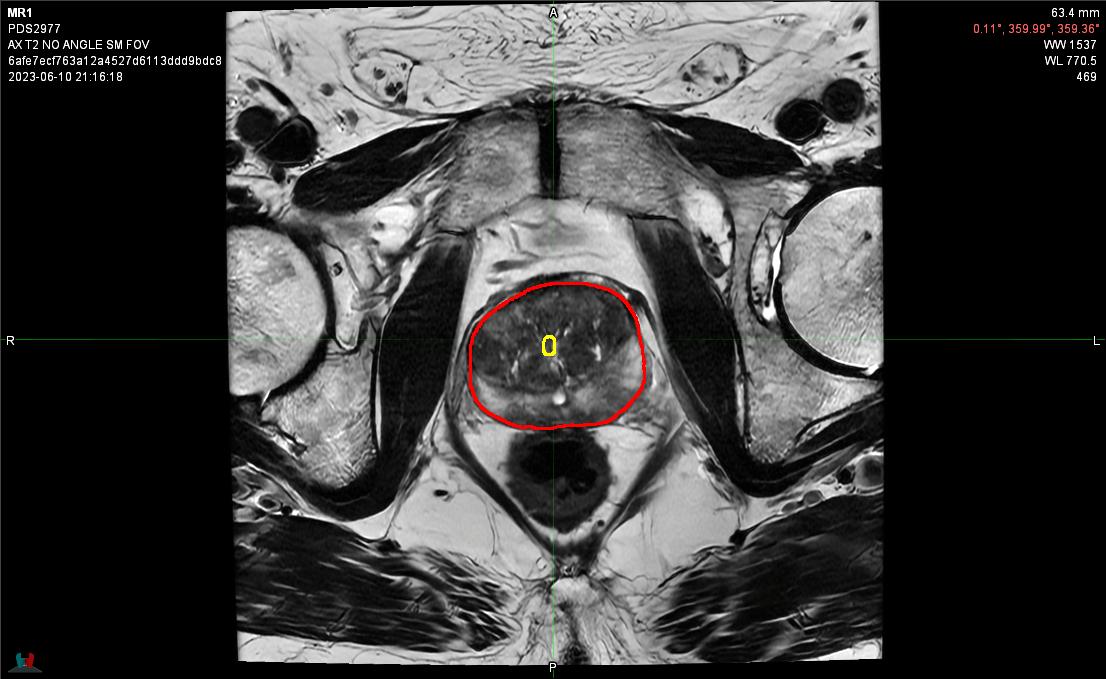  Axial View Slice 17 | 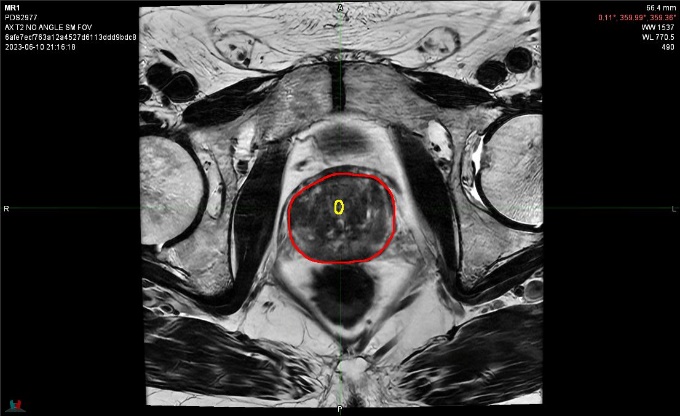  Axial View Slice 18 |
| 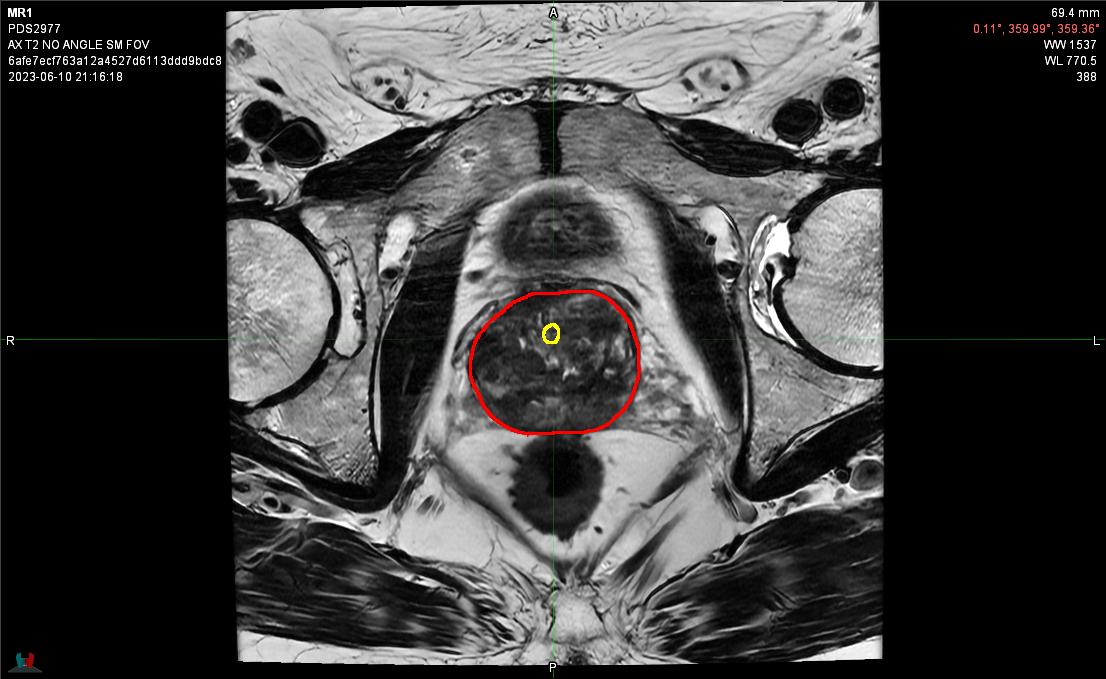  Axial View Slice 19 | 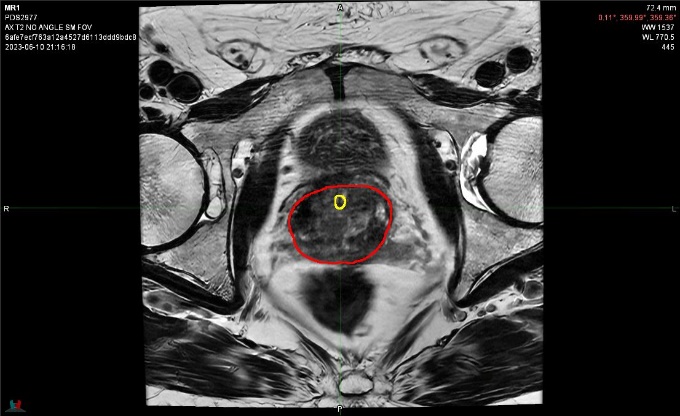  Axial View Slice 20 |
| 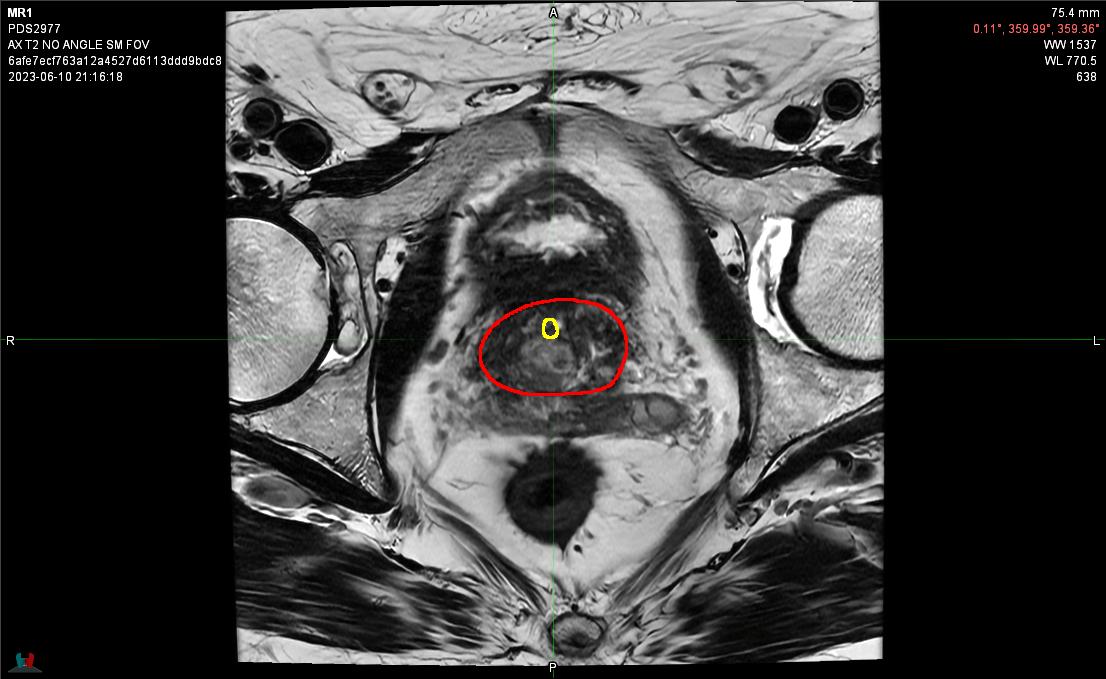  Axial View Slice 21 | 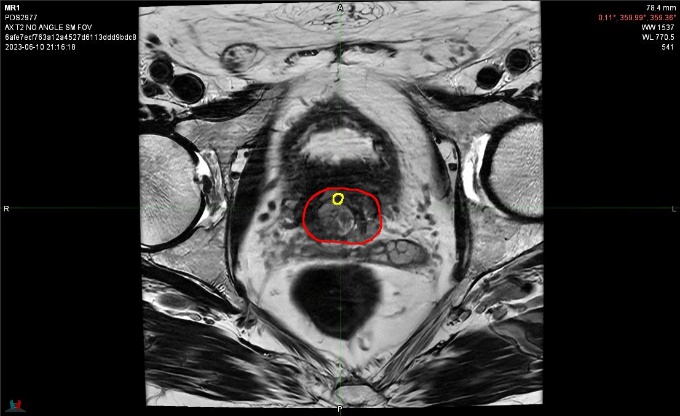  Axial View Slice 22 |
| 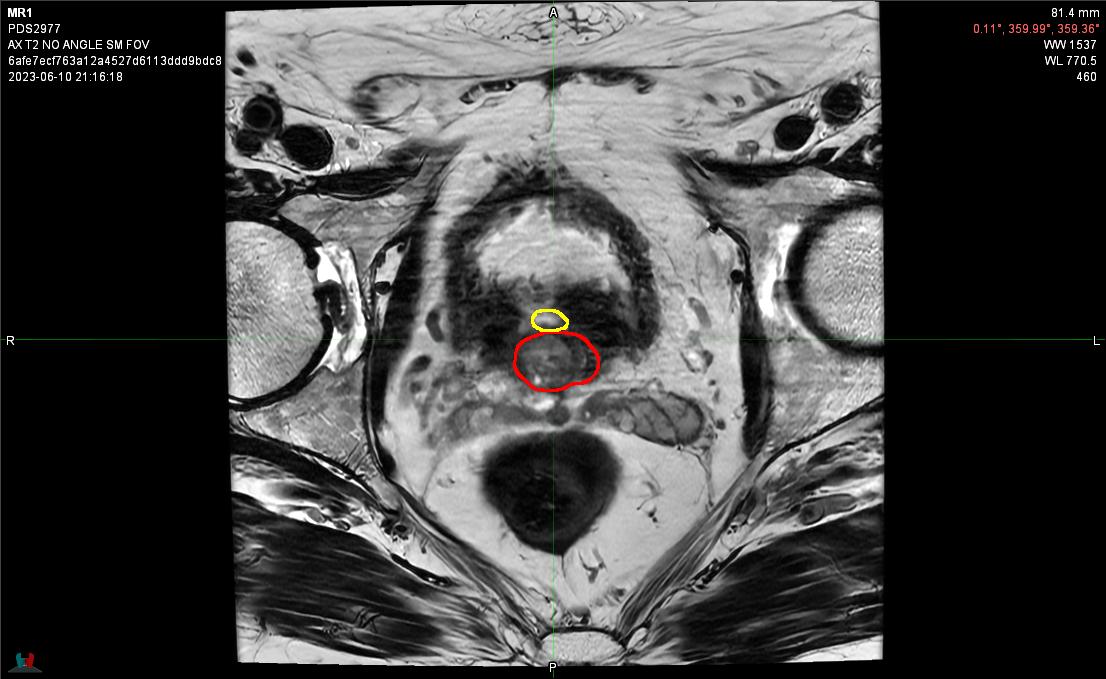  Axial View Slice 23 | 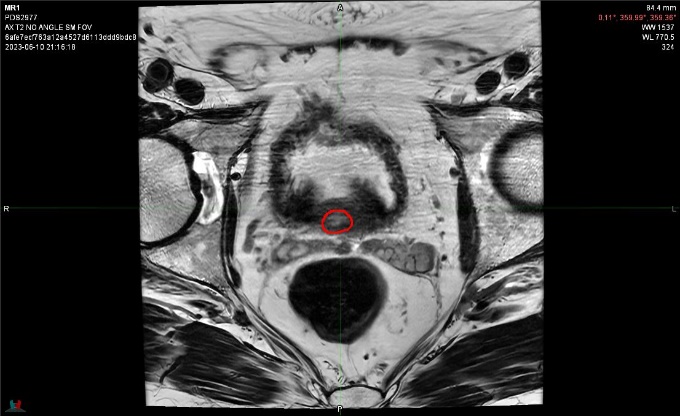  Axial View Slice 24 |
| 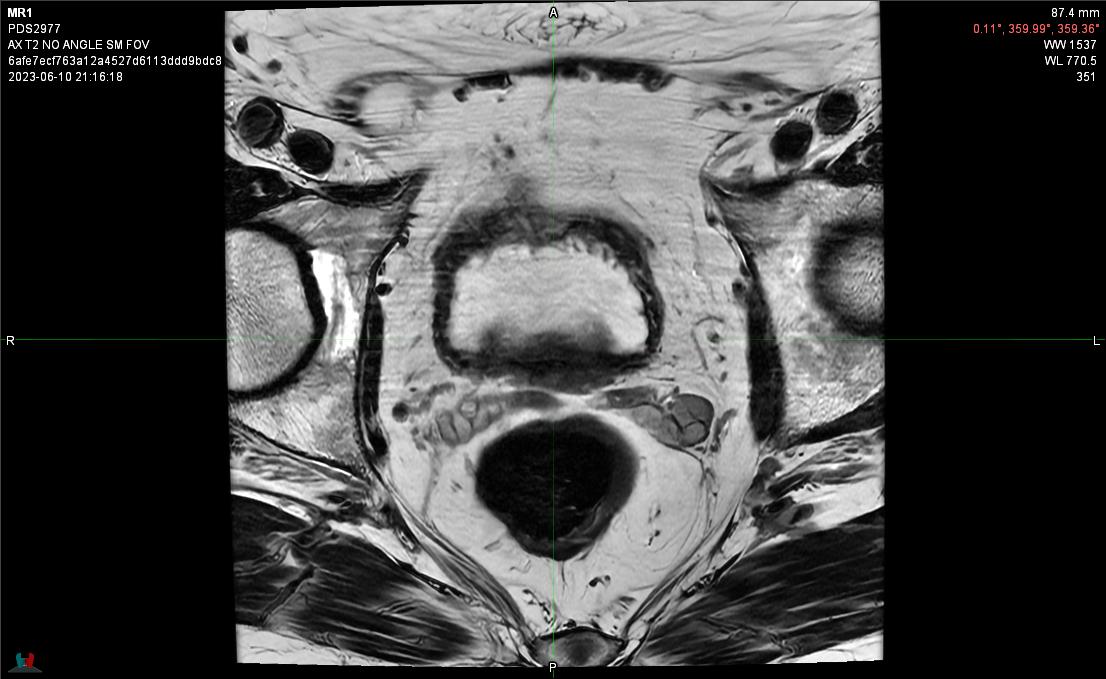  Axial View Slice 25 | 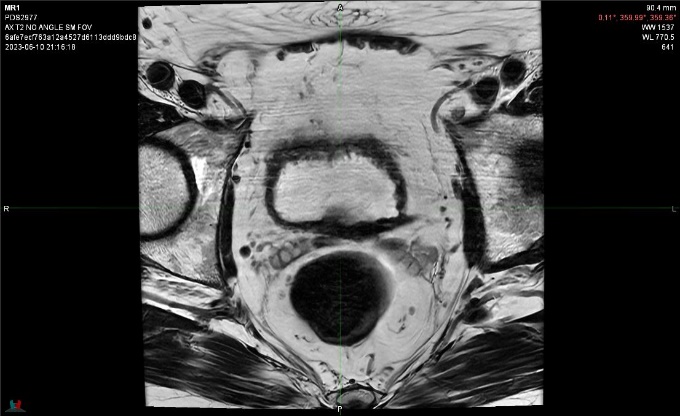  Axial View Slice 26 |
| 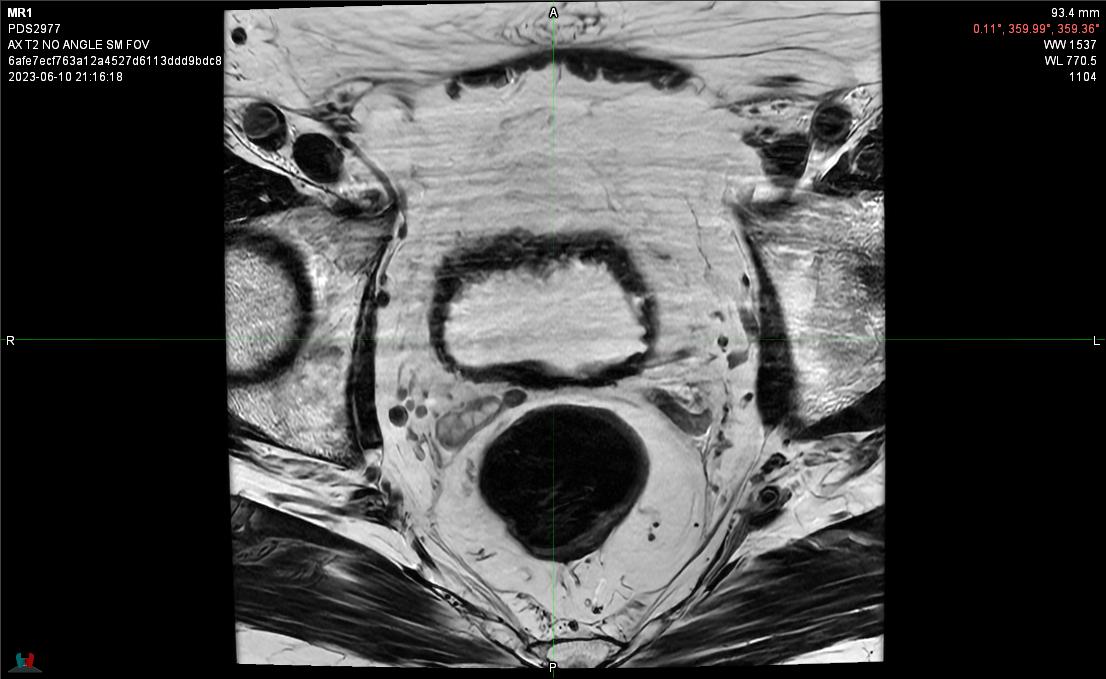  Axial View Slice 27 | 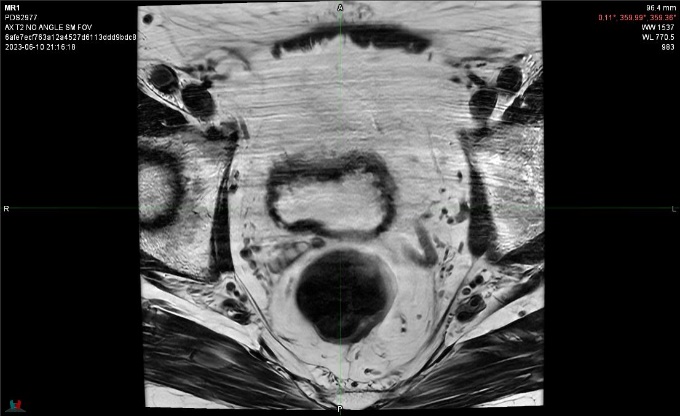  Axial View Slice 28 |
| 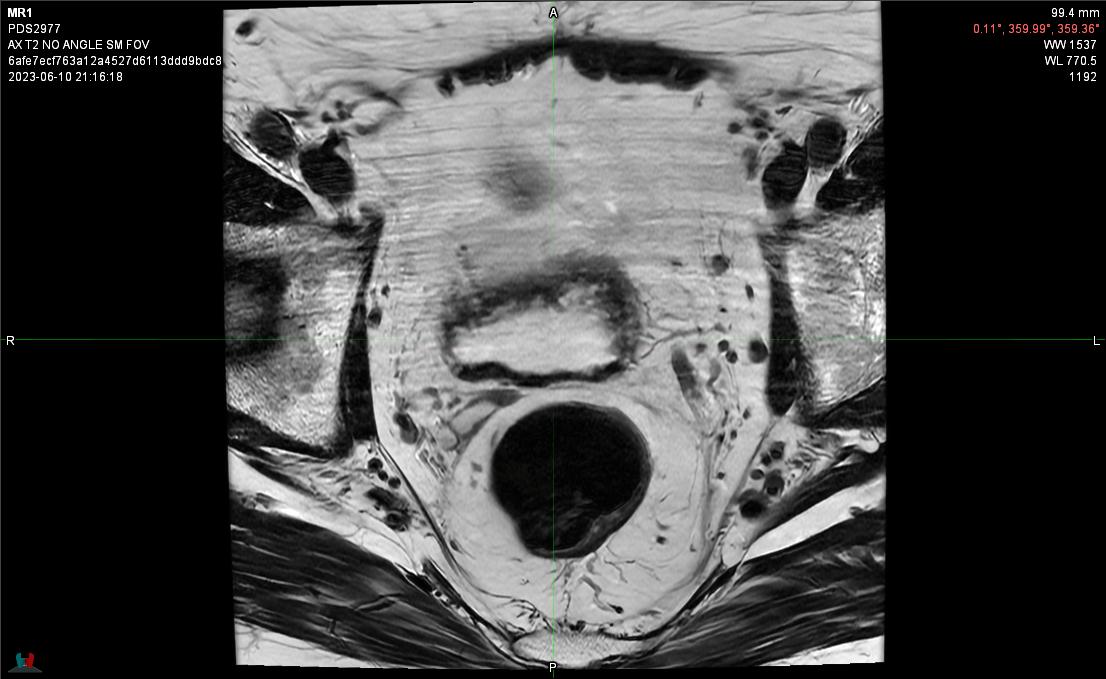  Axial View Slice 29 | 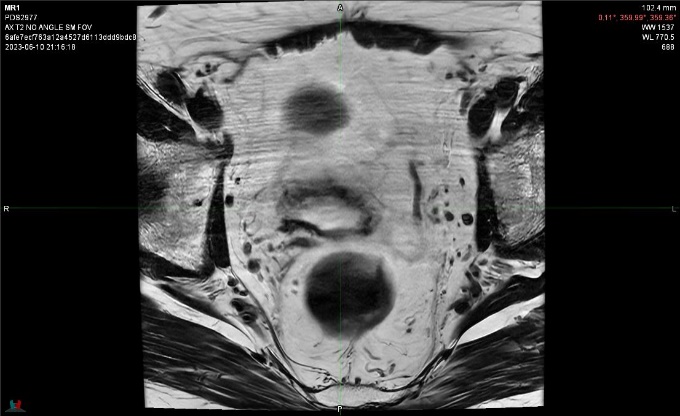  Axial View Slice 30 |
| 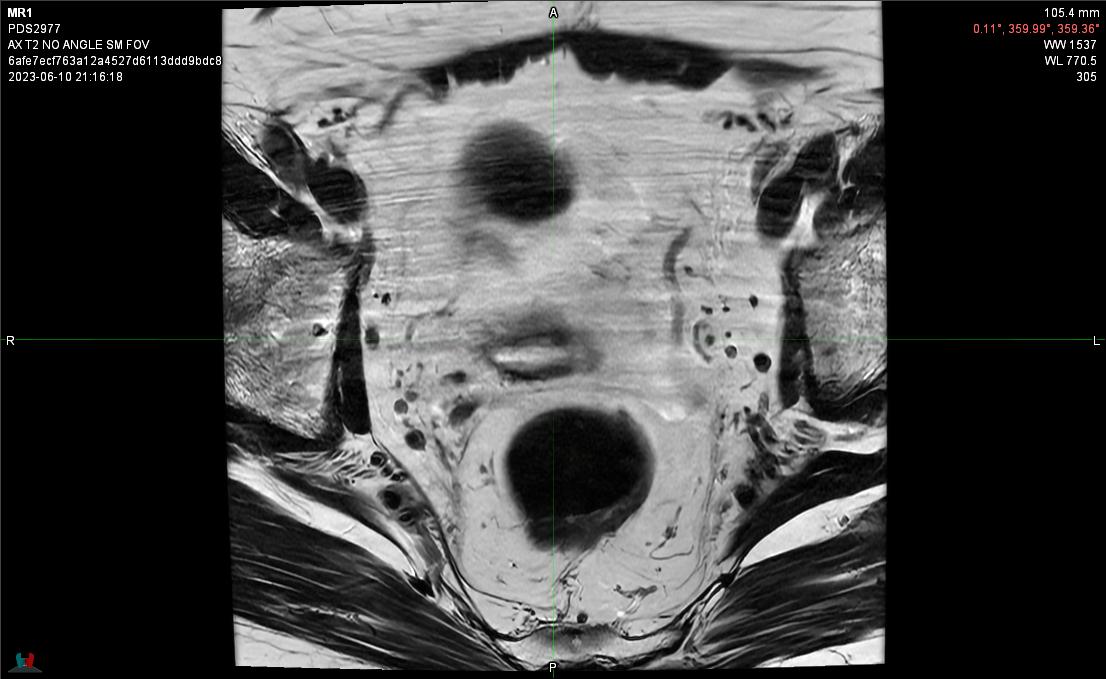  Axial View Slice 31 | 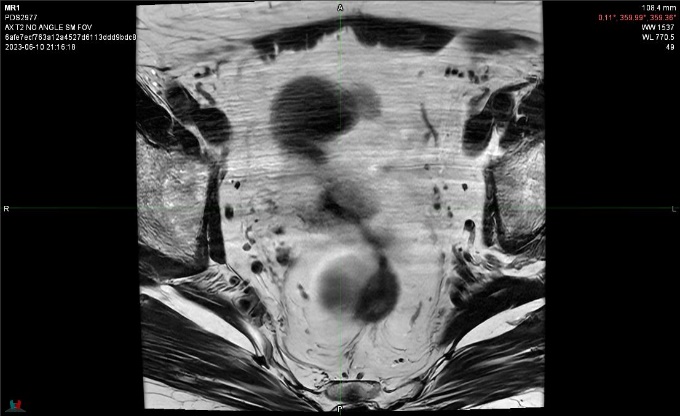  Axial View Slice 32 |
| 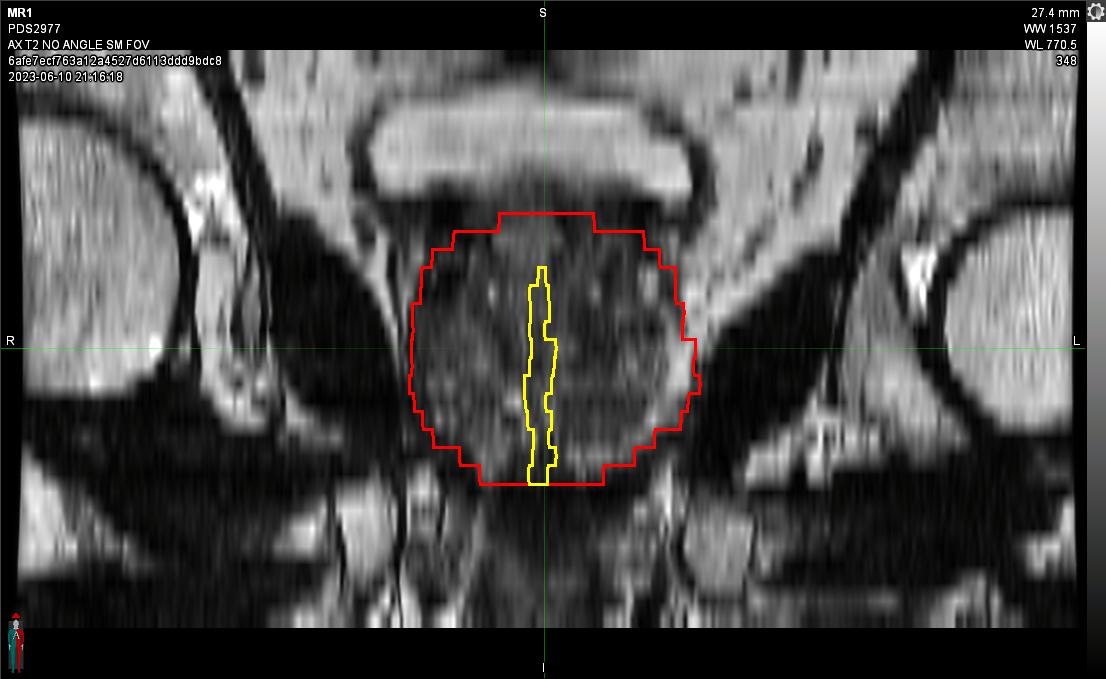Mid Coronal View | 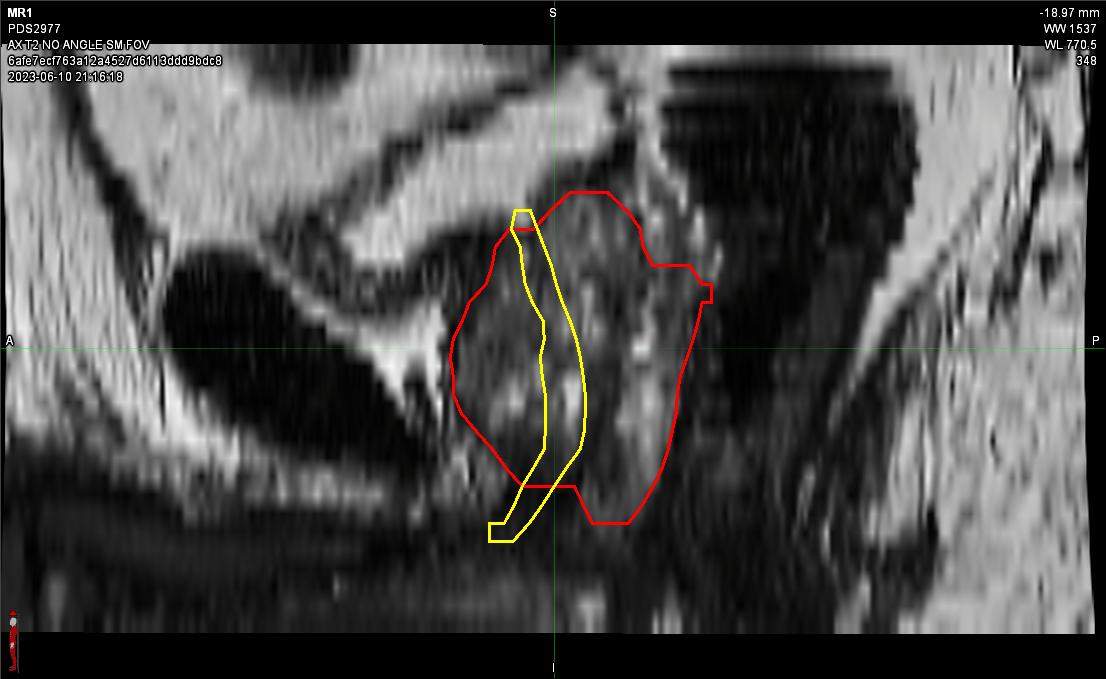Mid Sagittal View |

Supplementary Figure 2. Expert panel consensus contours for all slices from a representative patient case on axial *T2*-weighted slices on MRI. The contours in red are of the prostate, and the contours in yellow are of the urethra.
